# Factors associated with neuromusculoskeletal injury and disability in Navy and Marine Corps personnel

**DOI:** 10.1101/2022.06.09.22276233

**Authors:** John J. Fraser, Andrew J. MacGregor, Kenneth M. Fechner, Michael R. Galarneau

## Abstract

**Introduction:** Neuromusculoskeletal injuries (NMSKI) are ubiquitous in the military, which contribute to short- and long-term disability.

**Methods:** NMSKI, limited duty (LIMDU), and long-term disability episode counts in the US Navy (USN) and Marine Corps (USMC) from December 2016 to August 2021 were extracted from the Musculoskeletal Naval Epidemiological Surveillance Tool. NMSKI, LIMDU, and long-term disability episodes incidence were calculated. A hurdle negative binomial regression evaluated the association of body region, sex, age, rank, age by rank, and service branch on NMSKI, LIMDU, and long-term disability incidence.

**Results:** From December 2016 to August 2021, there were 2004196 NMSKI episodes (USN: 3285/1000 Sailors; USMC: 4418/1000 Marines), 16791 LIMDU episodes (USN: 32/1000 Sailors; USMC: 29/1000 Marines), and 2783 long-term disability episodes (USN: 5/1000 Sailors; USMC: 5/1000 Marines). There was a large-magnitude protective effect on NMSKI during the pandemic (RR, USN: 0.70; USMC: 0.75). Low back and ankle-foot were the most ubiquitous, primarily affecting female personnel, aged 25-44 years, senior enlisted, in the USMC. Shoulder, arm, pelvis-hip, and knee conditions had the greatest risk for disability, with female sex, enlisted ranks, ages 18-24 years, and service in the USMC the most salient factors.

**Discussion:** The significant protective effect during the pandemic was likely a function of reduced physical exposure and access to non-urgent care. Geographically accessible specialized care and resources aligned with communities with the greatest risk is needed for the timely prevention, assessment, and treatment of NMSKI.

**Conclusion:** Body region, sex, age, rank, and branch were salient factors for NMSKI.

## INTRODUCTION

Neuromusculoskeletal injuries (NMSKI) are ubiquitous in military personnel that threaten accomplishment of national security objectives.^1^ In best cases, these conditions preclude physical activity and limit a service member’s ability to function in their occupational specialties for only a finite period. More frequently, body system impairments and symptoms (such as pain) preclude function for protracted periods, which substantially affects quality of life, increases healthcare costs, and degrades mission readiness.^2^ The association between military activity and injury is likely bidirectional, since fluctuations in geopolitics and operational requirements influence the type and volume of exposure, environmental factors, and intrinsic factors associated with NMSKI in military servicemembers.^1^

There are multiple factors associated with NMSKI, including sex, occupation, age, rank, and the period of study. These factors, in addition to the suspected latent factors such as care-seeking behaviors, access to care, and the mode and duration of exposure, plausibly drive these outcomes in the military.^1^ This likely includes periods of reduced operational demand,^3–6^ something that occurred during 2020 COVID-19 pandemic as the US military shifted to a health protection posture required to mitigate the spread of this respiratory illness. A decline in NMSKI over time was observed in recent studies of the military, a finding that was attributed to the change in geopolitics, decline in combat operations, return to peacetime operations, and decreased exposure to hazards incurred during training and operations.^3–6^ This supposition can now be assessed in a more recent study epoch, where training and non-essential operations were halted to mitigate the spread of COVID-19.^7^

While military personnel frequently experience rapid recovery and return to occupational duties following NMSKI, others have protracted periods of activity limitation that preclude participation in organizational mission objectives.^2^ This is especially problematic considering the associated costs and implications for national security.^1,2^ Assessment of the factors that may be associated with NMSKI outcomes, short-term limited duty (LIMDU), and long-term disability (with medical attrition) would allow for greater precision for targeted prevention, optimization of healthcare delivery models, and identification of factors that may contribute to protracted care and long-term disablement. Therefore, the purpose of this study was to assess the effects of sex, age, rank, and service branch, and body region on the incidence of NMSKI, LIMDU, and long-term disability in the Department of the Navy (DoN), specifically members serving in the US Navy (USN) and US Marine Corps (USMC). A secondary aim was to evaluate the impact of the COVID-19 pandemic on the incidence of NMSKI.

## METHODS

A population-based epidemiological retrospective cohort study of all military members in the DoN was performed to assess body region, sex, age, rank, and service branch on the incidence of NMSKI, LIMDU, and long-term disability following these injuries from December 2016 to August 2021. The Strengthening the Reporting of Observational Studies in Epidemiology (STROBE) Statement was used to guide reporting.

NMSKI episodes, LIMDU, and long-term disability counts were extracted from the Musculoskeletal Naval Epidemiological Surveillance Tool [(MSK NEST), Navy Bureau of Medicine & Surgery, Falls Church, VA]. This surveillance tool leverages existing validated databases, such as the Military Health System Data Repository (MDR), Limited Duty Sailor and Marine Readiness Tracker System (LIMDU SMART), Disability Evaluation System (DES) and Defense Manpower Data Center Reporting System to report injury burden. NMSKI episodes within the MSK NEST were derived from healthcare encounters in the MDR and defined using the Army Public Health Center’s Taxonomy for Musculoskeletal Injuries based on an International Classification of Diagnosis, Tenth Revision (ICD-10) classification.^8^ Individuals with repeat visits for the same diagnosis in a single care episode were only counted once. The MSK NEST provides aggregated count data for NMSKI episodes by body region and de-identified patient characteristics that included sex; age range (18-24, 25-34, 35-44, _≥_45); rank [Junior Enlisted (E1-E5), Senior Enlisted (E6-E9), Junior Officers (O1-O3); Senior Officers (O4-O10)]; and service branch (USN, USMC) for all active and reserve personnel in the DoN. The database does not include any personal identifiable health information. This study was approved as non–human-subjects research by the Institutional Review Board at the U.S. Naval Health Research Center (NHRC.2022.0201.NHSR).

Since military end strength fluctuates annually due to attrition and recruitment of replacements,^9^ the population at risk was a dynamic cohort. Cumulative incidence of NMSKI was calculated and normalized to the subpopulation at risk (with consideration to sex, service branch, and rank) during the 4.75-year study epoch. Rates of LIMDU and long-term disability were calculated per 1000 episode-years for each subpopulation. In the evaluation of the effects of COVID-19, the mean NMSKI incidence was calculated for the pre-pandemic epoch (December 2016 to March 2020) and the during the pandemic (April 2020 to August 2021). Relative risk (RR) and 95% CI were calculated to assess the effects of the pandemic epoch.

A multivariable negative binomial regression was performed to evaluate the association of body region, sex, age, rank, age by rank, and branch of service on the incidence of NMSKI episodes, LIMDU, and long-term disability. Due to overdispersion of the data and the presence of excess zeros, a hurdle negative binomial model was employed over a Poisson or standard negative binomial model.^10^ In the hurdle negative binomial, the results are reported using calculations of the predictors regressed on count data (count model assessing the rate of the outcome), as well as a linked logistic regression (zero model which assess the probability of not having the outcome). The regression analyses were performed using the ‘MASS’ (version 7.3-54) and ‘PSCL’ (version 1.5) packages on R (version 3.5.1, The R Foundation for Statistical Computing, Vienna, Austria). Ankle-foot injuries served as the contrast in the assessment of body region. Female military personnel were selected as the reference group in the assessment of sex. The 18-24 age range was selected as the contrast for age. Junior enlisted personnel served the reference group for rank due to the greater disease and non-battle injuries in this group compared to commissioned officers.^11^ Finally, the USMC was the reference group for service branch. The level of significance was *p<*0.05 for all analyses. Statistical significance was evaluated using the convergence of odds ratio CIs that did not cross 1.00 and the *p*-value reported in the regression analysis.

## RESULTS

The incidence of NMSKI, NMSKI-related limited duty, and long-term disability stratified by body region, sex, age, rank, and service branch, in the USN, USMC, and DoN are reported in **Supplemental Tables 1-3**. From December 2016 to August 2021, there were 2004196 episodes of NMSKI (USN: 3285 episodes/1000 Sailors; USMC: 4418 episodes/1000 Marines), 16791 cases of LIMDU (USN: 32/1000 Sailors; USMC: 29/1000 Marines), and 2783 cases of long-term disability (USN: 5/1000 Sailors; USMC: 5/1000 Marines) in the DoN. **Figure 1** details the incidence of the NMSKI in the USN, USMC, and the total DoN during pre-pandemic and pandemic epochs. The pre-pandemic rate of NMSKI was 761.00 per 1000 person-years (95% CI: 733.41-788.08) in the USN; 1004.73 per 1000 person-years (95% CI: 973.03-1035.92) in the USMC; and 847.93 per 1000 person-years (95% CI: 818.81-876.55) in the total DoN. During the pandemic, the rate decreased to 534.65 per 1000 person-years (95% CI: 511.53-557.27) in the USN; 751.23 per 1000 person-years (95% CI: 723.82-778.18) in the USMC; and 608.96 per 1000 person-years (95% CI: 584.28-633.13) in the DoN, an indication of a large magnitude protective effect (RR, USN: 0.70; USMC: 0.75; DoN: 0.72). A large inflection in the rate of NMSKI was also noted in August 2021 during the final month of study.

**Figure 1.**
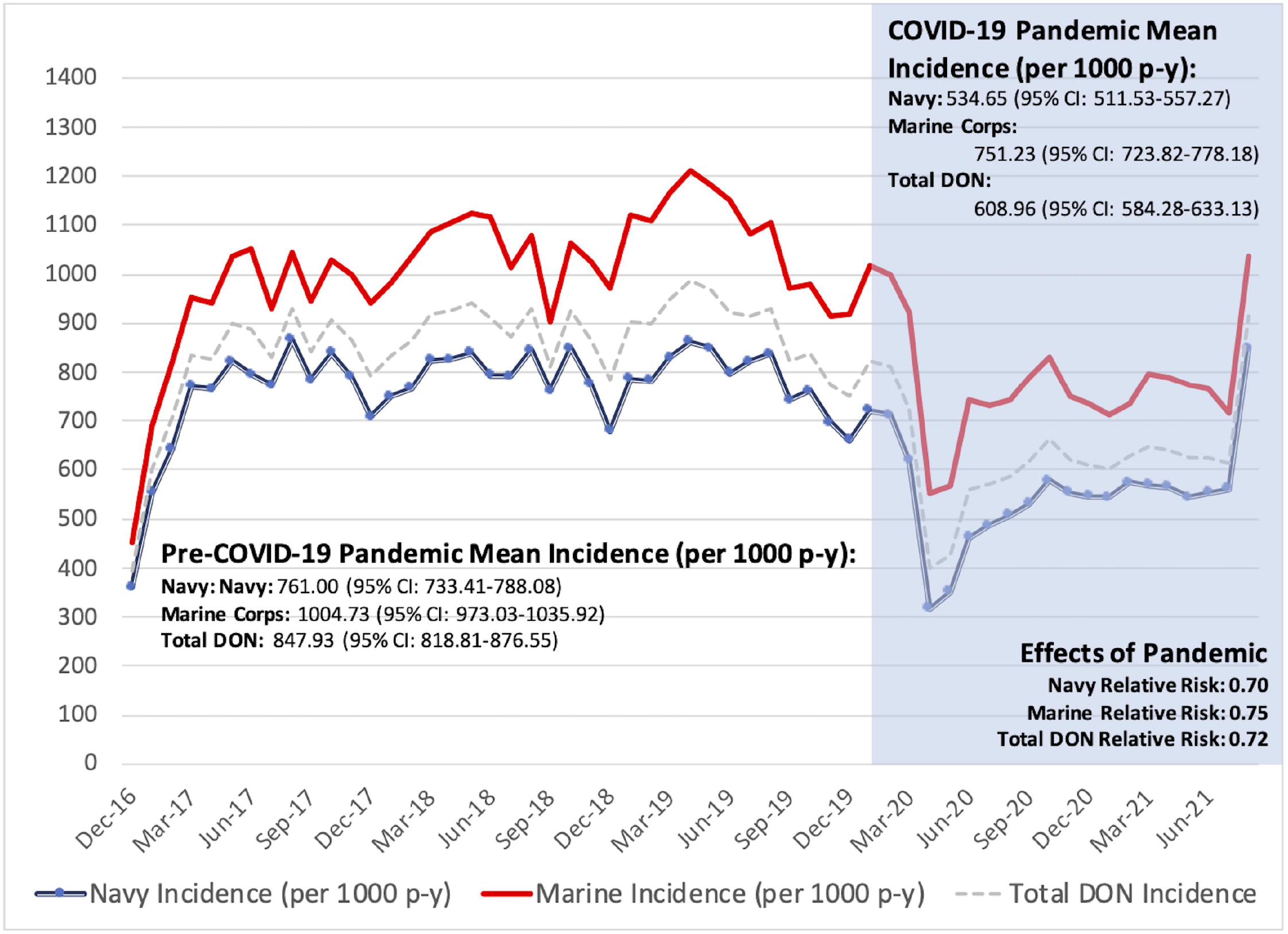
Effects of the COVID-19 pandemic on episodic neuromuscular injury incidence in the US Navy and US Marine Corps.

### NMSKI Factors

While incidence of ankle-foot injuries were second to only low back conditions across the DoN (**Table 1**), there were no significant differences in the incidence of these conditions when assessed in the individual branches. Sex was a salient factor, with male personnel observed to have reduced odds of experiencing at least one NMSKI (**Supplemental Table 4**). Once NMSKI occurred, males Sailors had lower rate ratios of injury compared to their female counterparts, with no significant difference in rates observed between male and female Marines. When contrasted to the 18–24-year-old group, there was a significantly lower odds of experiencing a NMSKI in Marines aged _≥_35, with no significant difference between ages in the USN. Once injury occurred, there was an increased incidence in Sailors aged 25-44 and a decreased rates in Marines aged _≥_45 group observed. Rank was an important factor, with Navy senior enlisted and officers and Marine junior officers demonstrating an increased odds of experiencing an injury compared to the junior enlisted. Once injury occurred, Navy senior enlisted had significantly higher rates and Marine junior officers had significantly lower rates of NMSKI than junior enlisted. The interactions of rank by body region and rank by age on injury incidence were non-significant or had observed singularities and were removed from the final model.

**Table 1.**
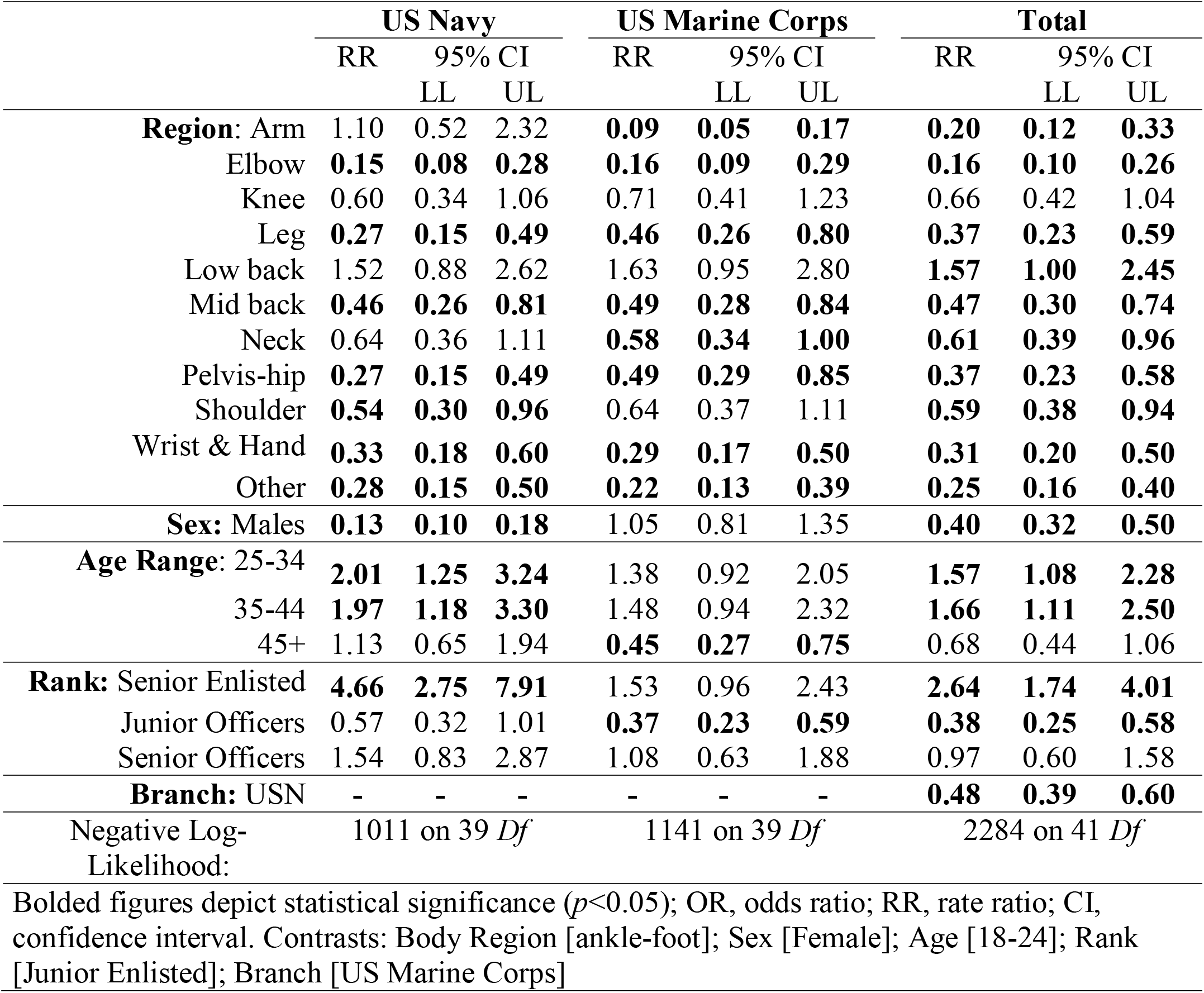
Results of the hurdle negative binomial regression assessing body region, sex, age, rank, and service branch on risk of neuromusculoskeletal injury episodes (adjusted for zero deflation) in US Navy (USN) and US Marine Corps personnel.

### Factors of NMSKI-related Limited Duty and Long-term Disability

**Table 2** details the assessment of LIMDU in the USN, USMC, and total DoN (complete results, to include the zero model of the hurdle negative binomial regression, are detailed in **Supplemental Table 5)**. In the USN, there was a significantly lower odds of incurring LIMDU resulting from arm, elbow, and mid back conditions. The rates of LIMDU were significantly higher in knee, pelvis-hip, and shoulder conditions and lower in the mid back compared to ankle-foot conditions in the USN and USMC. Only Marines had lower rates of LIMDU due to neck, wrist-hand, and other conditions compared to ankle-foot conditions. While male Sailors and Marines had significantly higher odds of incurring at least one NMSKI-related LIMDU, the rate was significantly lower in male Marines compared to their female counterparts. Sailors aged _≥_45 years and Marines _≥_35 years had significantly lower odds of being placed on LIMDU compared to personnel aged 18-24. Once placed in a limited-duty status, the rates were significantly lower in Sailors aged _≥_25 and Marines _≥_ 35 years compared to personnel aged 18-24. Senior enlisted members in the USN and USMC had significantly greater odds of being placed on LIMDU compared to junior enlisted, with Marine junior officers and senior Naval officers demonstrating significantly lower odds. All junior enlisted had significantly higher rates of LIMDU compared to all other ranks.

**Table 2.**
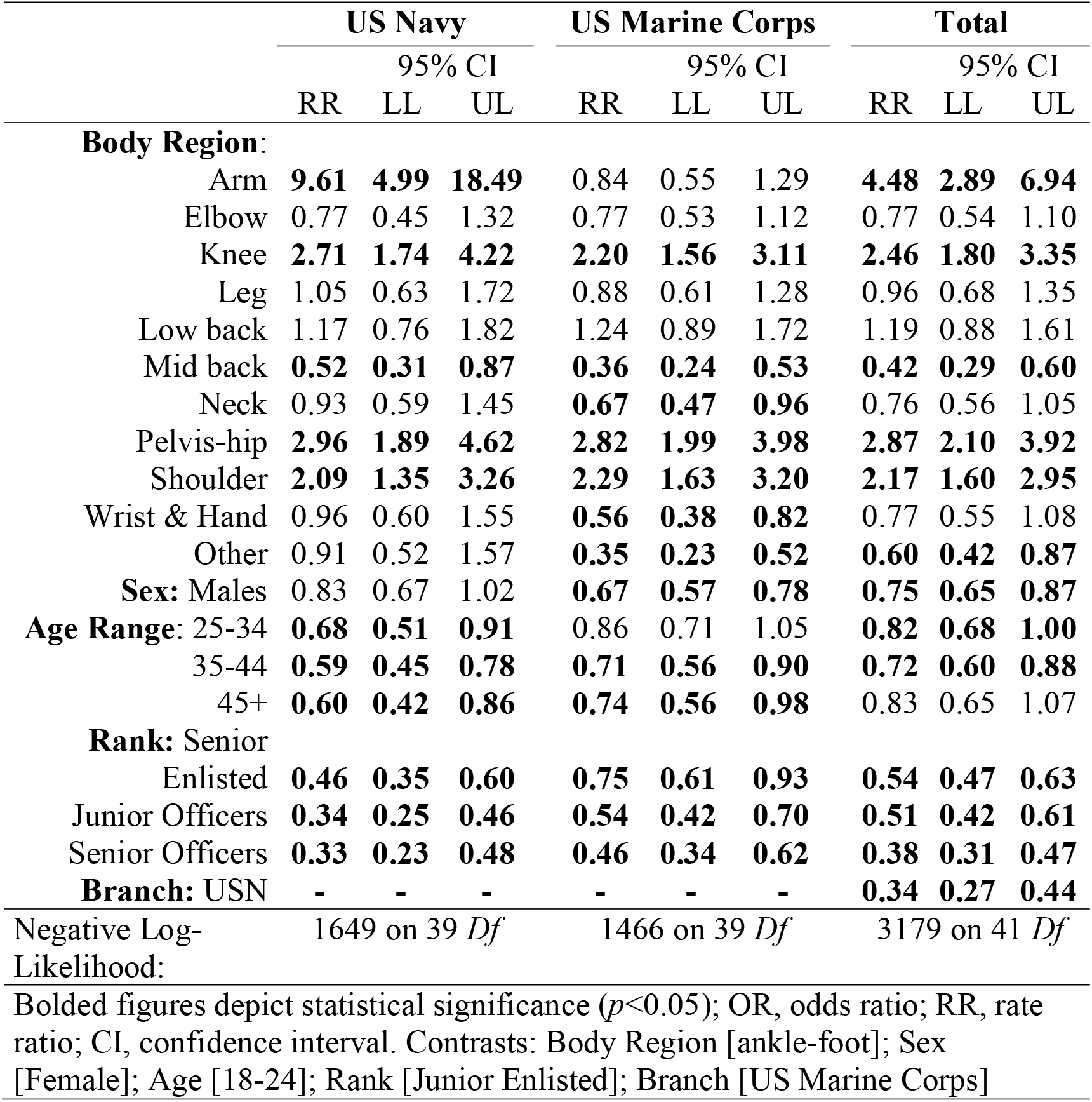
Results of the hurdle negative binomial regression assessing body region, sex, age, rank, and service branch on risk of neuromusculoskeletal injury-related episodes of limited duty (adjusted for zero deflation) in US Navy (USN) and US Marine Corps personnel.

**Table 3** details the results assessing body region, sex, age, rank, and service branch on long-term disability (complete results, to include the zero model of the hurdle negative binomial regression, are detailed in **Supplemental Table 6)**. Low back and other conditions, Marines aged _≥_35, and Marine officers had lower odds of experiencing long-term disability. Male sex was associated with increased odds of long-term disability. Sailors with arm, elbow, or knee conditions, Navy junior enlisted, and DoN personnel with pelvis-hip and shoulder conditions had the greatest incidence of long-term disability. Marines with neck conditions and Sailors aged _≥_45 had the lowest rates of long-term disability. When the results of the hurdle model were compared to the standard negative binomial regression in the assessment of adjusted injury risk, the zero-adjusted model outperformed the non-adjusted model in each of the outcomes assessed (**Supplemental Tables 7-9**).

**Table 3.**
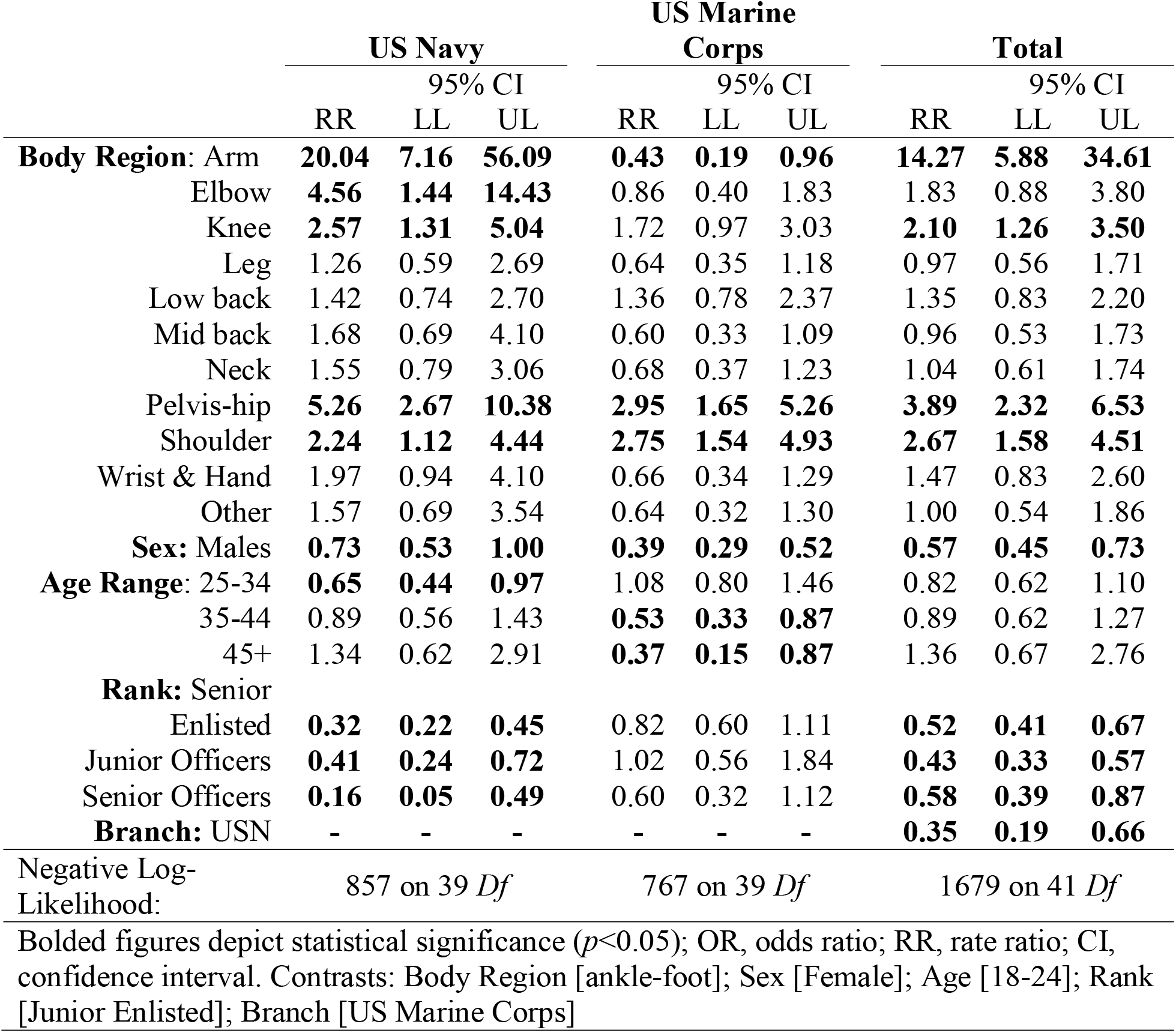
Results of the hurdle negative binomial regression assessing body region, sex, age, rank, and service branch on risk of neuromusculoskeletal injury-related episodes of long-term disability (adjusted for zero deflation) in US Navy (USN) and US Marine Corps personnel.

## DISCUSSION

The primary findings of this study were that during the COVID-19 pandemic, there was a significant protective effect for NMSKI that was likely a function of reduced physical exposure and access to non-urgent care. In the broader assessment of body region, sex, age, rank, and service branch on burden between 2016-2021, low back and ankle-foot were the most ubiquitous of all NMSKI in the DoN, primarily affecting female personnel, aged 25-44 years, senior enlisted, and Marines. Among the cases of LIMDU and long-term disability, female enlisted Marines, ages 18-24 years, with shoulder, arm, pelvis-hip, and knee conditions demonstrated the greatest risk. This is the first study to the authors’ knowledge to investigate the effects of the pandemic on NMSKI in the military, as well as the burden of NMSKI-related LIMDU and disability across the USN and USMC.

### NMSKI Burden

The protective effect observed during the COVID-19 pandemic is likely attributable to a shift in operations to only mission-critical tasks since infectious disease prophylaxis was prioritized to preserve force health protection. Medical operations similarly pivoted to urgent and emergent care, with the capability to provide routine care substantially curtailed. As such, reduced exposure to potentially injurious activities during military operations, combined with reduced healthcare utilization for NMSKI during the pandemic, are likely responsible for the reduction in NMSKI incidence. A similar phenomenon has been observed in NMSKI-related emergency department visits and surgical procedures in civilian hospitals.^12^ The rapid increase in incidence in the latter months of the study epoch corresponds with a period of resumption of regular operations, to include preparation for the fall physical readiness assessment required by the USN and USMC in 2021.

Occupational exposures differed between enlisted and officer personnel, with the former typically employed in vocational/technical fields and the latter responsible for administration and supervision of their subordinates. This may explain the lower NMSKI risk in the officer staff. Even within the enlisted ranks, exposures changes with increasing seniority during the transition to more supervisory and administrative roles. However, the substantially higher injury risk in senior enlisted personnel can be explained. It is highly conceivable that enlisted personnel who did not seek care for NMSKI earlier in their career may experience persistent symptoms, impairments, and activity limitation from chronic injury.^1^ It is also plausible that social determinants of care-seeking, such as stigma, social pressures, and access to NMSKI care improved as members advanced in rank. Differences in service cultures between branches may influence care-seeking determinants (e.g. mission over self; perceptions of weakness or NMSKI as self-limiting), access to care (geographic and time-based barriers), and exposure based on the specific mission requirements (with vastly different occupational demands and hazards) may also explain why the USMC had significantly larger risk compared to the USN.^1^ These suppositions warrant future investigation.

### Limited Duty and Long-term Disability

Short-term LIMDU and long-term disability were most common in conditions of the shoulder, arm, pelvis-hip, and knee. While impairments in these regions would certainly limit a personnel ability to push, pull, lift, kneel, squat, and negotiate terrain, ladders, and stairs during occupational tasks, these impairments would also affect a service member’s ability to perform on the annual physical fitness assessments used in the USN and USMC.^13,14^ These assessments encompass a timed running event (USN: 2.4 km, USMC: 4.8 km), a timed plank event [crunches are an optional alternative in USMC only], and an upper extremity endurance event (maximum pushups [pullups are an optional alternative in USMC only] in 2 minutes).^13,14^ Since performance of the biannual physical fitness assessment is a requirement in both the USN and USMC,^13,14^ it is not uncommon for clinicians to use performance on these tasks as criteria when making return to duty determination. Interestingly, personnel frequently have low confidence in their ability to perform occupational and physical readiness requirements following NMSKI, with persistent pain and reinjury common in personnel returned to full duty.^15^ The associations between neuromusculoskeletal impairment, perceptual and psychological factors, pain, and physical activity in personnel with injuries warrant further exploration.

Female sex, junior enlisted, aged 18-24 years, and Marines demonstrated the greatest risk of short-term LIMDU and long-term disability. Female sex has been found to be a salient factor for activity limitations and disability in veterans, but not active duty personnel, in a study of data derived from the US Census.^16^ This divergence may be attributed to changing roles of female personnel, with the current study epoch occurring following a policy change that opened all military occupations to women.^17^ The observation of higher disability in younger, junior enlisted members is likely a function of occupation-related physical demand, where senior enlisted and officer staff are able to continue performing administrative duty requirements while resting, rehabilitating, and recovering from NMSKI, whereas more junior personnel with highly physical occupations often cannot. The finding of lower rates of disability in older, more senior personnel are also likely attributed to the retirement benefit. In these groups, personnel are either emotionally or financially vested in the system (especially when they have more than 10 years of service toward the 20-year retirement) but are too young to retire. It is also plausible that the probability of experiencing a disabling condition following NMSKI that would preclude continued military service is substantially reduced due to lower exposure in the more senior personnel.

### Limitations

There are limitations to this study. First, this study relied on deidentified, aggregated data provided in the NEST database. While this allowed us to ascertain population-level burden of NMSKI incidence, short-term LIMDU, and long-term disability, the lack of individual-level data precluded the assessment of comorbidities and other intrinsic factors that may have contributed to the outcomes under study. The NEST database only provided data from the USN and USMC, which precluded assessment of the other branches of the US military and a comparison of service-related differences.

### Clinical Practice and Policy Implications

From a medical planning perspective, allocation of geographically accessible specialized care and resources needed for the timely prevention, assessment, and treatment of NMSKI should be aligned with communities that have the greatest risk.^1^ This evidence helps to prioritize allocation of staffing and supplies needed to address the populations with greatest risk of injury occurrence, short-term LIMDU, and long-term disability in order to optimize readiness. While global pandemics are infrequent and only occurring once every few generations, the release of biological agents by nations and non-state actors is a persistent threat to readiness of the armed forces. It is paramount that military leadership considers the physical and operational readiness of personnel during periods of restricted movement and reduced social interaction (e.g., pandemics).

## CONCLUSION

There was a significant protective effect during the COVID-19 pandemic that was likely a function of reduced physical exposure and access to non-urgent care. In the broader assessment, injuries to the low back and ankle-foot were the most ubiquitous of all NMSKI in the DoN, affecting a higher proportion of female personnel, those aged 25-44 years, senior enlisted, and members of the USMC. Among the cases of LIMDU and long-term disability, conditions of the shoulder and arm in the upper quarter and pelvis-hip and knee in the lower quarter had the greatest burden, with personnel who were female, enlisted, ages 18-24 years, and serving in the Marine Corps having the greatest risk.

## Data Availability

All data produced in the present work are contained in the manuscript

