## Supplemental Tables 1-9 for "Factors associated with neuromusculoskeletal injury and disability in Navy and Marine Corps personnel"

**Supplemental Table 1. Incidence of neuromusculoskeletal injury (per 1000 person-years) in the US Marine Corps and US Navy from December 2016 to August 2021.**

| | US Marine Corps | | | | | | | | | | Mean Total | US Navy | | | | | | | | | | Mean Total |
|---|---|---|---|---|---|---|---|---|---|---|---|---|---|---|---|---|---|---|---|---|---|---|
| | Females | | | | Mean | Males | | | | Mean | | Females | | | | Mean | Males | | | | Mean | |
| | 18-24 | 25-34 | 35-44 | 45+ | | 18-24 | 25-34 | 35-44 | 45+ | | | 18-24 | 25-34 | 35-44 | 45+ | | 18-24 | 25-34 | 35-44 | 45+ | | |
| **Ankle Foot** | **50.04** | **38.45** | **35.92** | **14.00** | **33.58** | **28.47** | **50.18** | **50.98** | **13.67** | **36.31** | **34.95** | **24.77** | **95.40** | **92.15** | **27.78** | **62.38** | **1.67** | **8.35** | **6.95** | **5.33** | **5.84** | **34.11** |
| Junior Enlisted | 111.66 | 12.58 | 0.02 | 0.00 | 31.07 | 43.46 | 5.99 | 0.02 | 0.00 | 12.37 | 21.72 | 22.80 | 10.40 | 0.79 | 0.01 | 8.50 | 1.98 | 1.19 | 0.11 | 0.00 | 0.82 | 4.66 |
| Senior Enlisted | 22.42 | 68.52 | 35.72 | 5.33 | 33.00 | 35.03 | 160.20 | 155.64 | 29.03 | 94.97 | 63.99 | 48.01 | 326.34 | 285.17 | 57.13 | 179.16 | 2.55 | 27.14 | 13.50 | 8.95 | 13.04 | 96.10 |
| Junior Officer | 16.05 | 50.06 | 7.06 | 3.85 | 19.26 | 6.92 | 31.37 | 13.33 | 0.00 | 12.90 | 16.08 | 3.51 | 36.34 | 19.82 | 13.32 | 18.25 | 0.49 | 4.31 | 3.96 | 3.72 | 3.12 | 10.68 |
| Senior Officer | | 22.65 | 100.89 | 46.84 | 56.79 | | 3.14 | 34.93 | 25.65 | 21.24 | 39.02 | | 8.49 | 62.83 | 40.68 | 37.33 | | 0.76 | 10.24 | 8.66 | 6.56 | 21.95 |
| **Arm** | **3.45** | **3.08** | **4.37** | **2.10** | **3.24** | **2.99** | **6.61** | **7.40** | **1.82** | **4.82** | **4.03** | **2.92** | **8.96** | **8.35** | **2.84** | **5.96** | **18.73** | **1.08** | **0.86** | **0.65** | **4.44** | **5.20** |
| Junior Enlisted | 6.49 | 0.95 | 0.00 | 0.00 | 1.86 | 4.06 | 0.63 | 0.00 | 0.00 | 1.17 | 1.52 | 1.91 | 1.03 | 0.10 | 0.00 | 0.76 | 55.75 | 0.14 | 0.01 | 0.00 | 13.98 | 7.37 |
| Senior Enlisted | 2.08 | 6.81 | 3.47 | 0.74 | 3.28 | 4.48 | 20.89 | 22.18 | 3.51 | 12.77 | 8.02 | 6.41 | 31.48 | 26.32 | 5.84 | 17.51 | 0.37 | 3.54 | 1.62 | 1.10 | 1.66 | 9.59 |
| Junior Officer | 1.77 | 3.53 | 1.12 | 0.96 | 1.85 | 0.43 | 4.34 | 2.02 | 0.00 | 1.70 | 1.77 | 0.45 | 2.95 | 1.74 | 1.10 | 1.56 | 0.05 | 0.54 | 0.52 | 0.47 | 0.39 | 0.98 |
| Senior Officer | | 1.03 | 12.87 | 6.69 | 6.86 | | 0.60 | 5.41 | 3.78 | 3.26 | 5.06 | | 0.37 | 5.23 | 4.43 | 3.34 | | 0.11 | 1.29 | 1.04 | 0.81 | 2.08 |
| **Elbow** | **2.74** | **3.84** | **5.61** | **3.28** | **3.94** | **3.72** | **10.43** | **17.22** | **5.28** | **9.53** | **6.74** | **2.33** | **8.95** | **12.54** | **5.52** | **7.67** | **0.22** | **1.60** | **2.07** | **1.68** | **1.47** | **4.57** |
| Junior Enlisted | 5.01 | 0.97 | 0.02 | 0.00 | 1.50 | 4.34 | 0.85 | 0.01 | 0.00 | 1.30 | 1.40 | 1.32 | 0.95 | 0.09 | 0.00 | 0.59 | 0.20 | 0.17 | 0.02 | 0.00 | 0.10 | 0.34 |
| Senior Enlisted | 2.08 | 8.90 | 7.09 | 0.74 | 4.70 | 5.77 | 32.49 | 50.87 | 10.71 | 24.96 | 14.83 | 5.42 | 30.96 | 36.90 | 10.89 | 21.04 | 0.38 | 5.13 | 3.67 | 2.53 | 2.93 | 11.99 |
| Junior Officer | 1.12 | 4.97 | 1.44 | 2.09 | 2.41 | 1.05 | 7.44 | 4.73 | 0.00 | 3.30 | 2.86 | 0.25 | 3.09 | 3.34 | 2.67 | 2.34 | 0.08 | 0.96 | 1.21 | 1.34 | 0.90 | 1.62 |
| Senior Officer | | 0.51 | 13.90 | 10.29 | 8.24 | | 0.95 | 13.27 | 10.43 | 8.22 | 8.23 | | 0.80 | 9.85 | 8.49 | 6.38 | | 0.15 | 3.38 | 2.87 | 2.14 | 4.26 |
| **Knee** | **36.23** | **26.43** | **23.41** | **6.89** | **22.37** | **29.73** | **44.74** | **42.97** | **10.03** | **32.01** | **27.19** | **17.76** | **57.73** | **48.80** | **15.73** | **36.15** | **1.54** | **6.61** | **4.27** | **3.45** | **4.13** | **20.14** |
| Junior Enlisted | 83.01 | 8.57 | 0.09 | 0.00 | 22.91 | 44.92 | 5.97 | 0.03 | 0.00 | 12.73 | 17.82 | 18.02 | 6.92 | 0.52 | 0.00 | 6.36 | 1.83 | 1.02 | 0.08 | 0.00 | 0.73 | 3.55 |
| Senior Enlisted | 19.60 | 50.04 | 24.05 | 3.10 | 24.20 | 40.54 | 145.72 | 132.49 | 21.31 | 85.02 | 54.61 | 32.84 | 201.14 | 155.43 | 35.13 | 106.14 | 2.45 | 21.68 | 7.79 | 5.84 | 9.44 | 57.79 |
| Junior Officer | 6.10 | 30.65 | 7.22 | 1.28 | 11.31 | 3.74 | 24.88 | 12.15 | 0.00 | 10.19 | 10.75 | 2.42 | 18.24 | 9.13 | 7.00 | 9.20 | 0.35 | 3.27 | 2.80 | 2.31 | 2.18 | 5.69 |
| Senior Officer | | 16.47 | 62.28 | 23.16 | 33.97 | | 2.40 | 27.21 | 18.80 | 16.14 | 25.06 | | 4.62 | 30.09 | 20.80 | 18.50 | | 0.47 | 6.42 | 5.66 | 4.18 | 11.34 |
| **Leg** | **32.24** | **16.32** | **11.19** | **2.85** | **14.54** | **18.63** | **24.49** | **20.61** | **4.85** | **17.05** | **15.79** | **11.25** | **27.23** | **20.97** | **6.60** | **16.86** | **1.02** | **3.30** | **1.93** | **1.46** | **1.99** | **9.43** |
| Junior Enlisted | 73.25 | 6.06 | 0.04 | 0.00 | 19.84 | 30.91 | 3.42 | 0.02 | 0.00 | 8.59 | 14.21 | 14.85 | 3.88 | 0.36 | 0.01 | 4.78 | 1.39 | 0.59 | 0.05 | 0.00 | 0.51 | 2.64 |
| Senior Enlisted | 11.91 | 26.13 | 10.47 | 1.62 | 12.53 | 20.02 | 76.86 | 60.84 | 8.84 | 41.64 | 27.09 | 17.30 | 93.82 | 65.05 | 11.83 | 47.00 | 1.40 | 10.45 | 3.11 | 2.10 | 4.27 | 25.63 |
| Junior Officer | 11.55 | 25.35 | 3.37 | 0.00 | 10.07 | 4.95 | 15.82 | 6.28 | 0.00 | 6.76 | 8.42 | 1.60 | 10.06 | 3.99 | 3.37 | 4.76 | 0.26 | 1.85 | 1.33 | 1.10 | 1.13 | 2.94 |
| Senior Officer | | 7.72 | 30.88 | 9.78 | 16.13 | | 1.86 | 15.30 | 10.58 | 9.25 | 12.69 | | 1.17 | 14.46 | 11.20 | 8.94 | | 0.31 | 3.23 | 2.64 | 2.06 | 5.50 |
| **Low back** | **67.89** | **64.80** | **62.30** | **12.47** | **50.80** | **58.17** | **111.47** | **94.61** | **20.15** | **71.96** | **61.38** | **48.21** | **153.31** | **111.27** | **29.68** | **88.11** | **3.68** | **17.95** | **11.36** | **6.98** | **10.42** | **49.26** |
| Junior Enlisted | 139.24 | 16.42 | 0.09 | 0.00 | 38.94 | 73.27 | 10.95 | 0.07 | 0.01 | 21.08 | 30.01 | 37.15 | 17.05 | 1.16 | 0.00 | 13.84 | 3.77 | 2.35 | 0.18 | 0.00 | 1.57 | 7.71 |
| Senior Enlisted | 49.02 | 121.25 | 52.21 | 5.28 | 56.94 | 93.97 | 352.88 | 288.33 | 42.87 | 194.51 | 125.73 | 101.74 | 531.35 | 342.03 | 58.27 | 258.35 | 6.48 | 58.48 | 23.01 | 11.93 | 24.97 | 141.66 |
| Junior Officer | 15.40 | 88.58 | 18.29 | 2.41 | 31.17 | 7.25 | 75.04 | 26.75 | 0.02 | 27.27 | 29.22 | 5.73 | 51.75 | 26.00 | 15.09 | 24.64 | 0.80 | 9.48 | 6.28 | 4.93 | 5.37 | 15.01 |
| Senior Officer | | 32.94 | 178.61 | 42.21 | 84.59 | | 7.03 | 63.29 | 37.71 | 36.01 | 60.30 | | 13.11 | 75.88 | 45.35 | 44.78 | | 1.51 | 15.98 | 11.06 | 9.52 | 27.15 |
| **Mid back** | **23.70** | **23.67** | **19.16** | **2.87** | **16.92** | **17.88** | **32.80** | **24.13** | **4.21** | **19.88** | **18.40** | **20.31** | **60.86** | **36.20** | **7.39** | **31.91** | **1.33** | **5.78** | **3.11** | **1.56** | **3.05** | **17.48** |
| Junior Enlisted | 48.70 | 5.59 | 0.00 | 0.00 | 13.57 | 23.85 | 3.19 | 0.02 | 0.00 | 6.77 | 10.17 | 16.50 | 7.23 | 0.37 | 0.00 | 6.03 | 1.41 | 0.78 | 0.05 | 0.00 | 0.56 | 3.29 |
| Senior Enlisted | 17.10 | 43.50 | 14.59 | 1.20 | 19.10 | 27.38 | 103.53 | 71.46 | 8.53 | 52.73 | 35.91 | 42.48 | 210.53 | 108.36 | 14.07 | 93.86 | 2.27 | 18.45 | 5.87 | 2.46 | 7.26 | 50.56 |
| Junior Officer | 5.30 | 33.22 | 3.37 | 0.48 | 10.59 | 2.41 | 22.23 | 7.44 | 0.00 | 8.02 | 9.31 | 1.94 | 20.80 | 7.76 | 3.06 | 8.39 | 0.30 | 3.30 | 1.83 | 1.15 | 1.64 | 5.02 |
| Senior Officer | | 12.35 | 58.68 | 9.78 | 26.94 | | 2.26 | 17.62 | 8.32 | 9.40 | 18.17 | | 4.86 | 28.31 | 12.43 | 15.20 | | 0.61 | 4.70 | 2.62 | 2.64 | 8.92 |
| **Neck** | **14.89** | **26.50** | **32.95** | **8.34** | **21.06** | **9.92** | **29.66** | **35.00** | **9.25** | **21.69** | **21.38** | **16.81** | **66.70** | **61.97** | **18.28** | **42.55** | **0.97** | **5.44** | **4.70** | **3.18** | **3.75** | **23.15** |
| Junior Enlisted | 27.81 | 4.55 | 0.02 | 0.00 | 8.10 | 11.44 | 2.14 | 0.02 | 0.00 | 3.40 | 5.75 | 12.37 | 6.23 | 0.51 | 0.01 | 4.78 | 0.95 | 0.67 | 0.06 | 0.00 | 0.42 | 2.60 |
| Senior Enlisted | 12.37 | 44.99 | 23.49 | 2.13 | 20.74 | 16.97 | 90.37 | 103.38 | 18.58 | 57.33 | 39.04 | 35.44 | 225.07 | 178.42 | 33.10 | 118.01 | 1.70 | 16.96 | 8.47 | 4.87 | 8.00 | 63.00 |
| Junior Officer | 4.49 | 39.47 | 9.47 | 2.41 | 13.96 | 1.35 | 23.39 | 11.40 | 0.00 | 9.03 | 11.50 | 2.61 | 28.84 | 15.46 | 9.16 | 14.02 | 0.27 | 3.48 | 2.79 | 2.46 | 2.25 | 8.13 |
| Senior Officer | | 16.99 | 98.83 | 28.83 | 48.21 | | 2.75 | 25.20 | 18.42 | 15.46 | 31.84 | | 6.65 | 53.48 | 30.83 | 30.32 | | 0.66 | 7.47 | 5.38 | 4.51 | 17.41 |
| **Other** | **12.84** | **10.36** | **12.33** | **1.89** | **9.12** | **8.45** | **14.01** | **10.20** | **2.42** | **8.79** | **8.96** | **10.47** | **33.97** | **26.40** | **7.62** | **20.22** | **0.73** | **2.62** | **1.50** | **1.03** | **1.52** | **10.87** |
| Junior Enlisted | 27.08 | 3.28 | 0.06 | 0.00 | 7.61 | 11.54 | 1.40 | 0.01 | 0.00 | 3.24 | 5.42 | 9.48 | 4.01 | 0.23 | 0.00 | 3.43 | 0.88 | 0.40 | 0.03 | 0.00 | 0.33 | 1.88 |
| Senior Enlisted | 7.74 | 23.54 | 9.78 | 0.74 | 10.45 | 11.46 | 44.10 | 31.20 | 4.61 | 22.84 | 16.64 | 20.33 | 117.59 | 78.86 | 13.76 | 57.64 | 1.13 | 8.49 | 2.98 | 1.72 | 3.58 | 30.61 |
| Junior Officer | 3.69 | 12.03 | 4.49 | 0.64 | 5.22 | 2.36 | 8.67 | 2.75 | 0.00 | 3.44 | 4.33 | 1.60 | 12.00 | 6.27 | 3.60 | 5.87 | 0.19 | 1.38 | 0.92 | 0.70 | 0.80 | 3.33 |
| Senior Officer | | 2.57 | 35.00 | 6.18 | 14.58 | | 1.86 | 6.83 | 5.07 | 4.59 | 9.59 | | 2.28 | 20.25 | 13.11 | 11.88 | | 0.22 | 2.09 | 1.69 | 1.34 | 6.61 |
| **Pelvis Hip** | **52.22** | **21.89** | **19.02** | **7.16** | **23.26** | **7.46** | **13.68** | **15.42** | **4.42** | **10.43** | **16.85** | **7.94** | **26.10** | **26.39** | **9.15** | **18.02** | **0.34** | **1.63** | **1.37** | **1.25** | **1.20** | **9.61** |
| Junior Enlisted | 129.25 | 9.52 | 0.00 | 0.00 | 34.69 | 11.29 | 1.58 | 0.01 | 0.00 | 3.22 | 18.96 | 7.64 | 2.85 | 0.18 | 0.00 | 2.67 | 0.38 | 0.23 | 0.02 | 0.00 | 0.16 | 1.41 |
| Senior Enlisted | 18.44 | 42.07 | 18.21 | 1.95 | 20.17 | 9.68 | 42.06 | 45.13 | 8.96 | 26.45 | 23.31 | 15.17 | 86.32 | 75.11 | 14.07 | 47.67 | 0.52 | 4.99 | 2.26 | 1.69 | 2.36 | 25.02 |

| | | | | | | | | | | | | | | | | | | | | | | |
|---|---|---|---|---|---|---|---|---|---|---|---|---|---|---|---|---|---|---|---|---|---|---|
| Junior Officer | 8.99 | 28.24 | 6.42 | 0.96 | 11.15 | 1.42 | 9.89 | 4.79 | 0.02 | 4.03 | 7.59 | 1.01 | 11.92 | 6.80 | 5.28 | 6.25 | 0.11 | 1.08 | 0.87 | 1.03 | 0.77 | 3.51 |
| Senior Officer | | 7.72 | 51.47 | 25.74 | 28.31 | | 1.19 | 11.74 | 8.71 | 7.21 | 17.76 | | 3.32 | 23.45 | 17.23 | 14.67 | | 0.23 | 2.33 | 2.27 | 1.61 | 8.14 |
| **Shoulder** | **16.05** | **22.09** | **19.44** | **7.52** | **16.29** | **20.05** | **44.79** | **48.98** | **12.68** | **32.40** | **24.34** | **9.82** | **37.23** | **37.85** | **14.03** | **25.73** | **1.19** | **6.30** | **4.91** | **4.12** | **4.32** | **15.03** |
| Junior Enlisted | 32.39 | 4.08 | 0.02 | 0.00 | 9.12 | 23.52 | 3.80 | 0.02 | 0.00 | 6.84 | 7.98 | 7.36 | 3.75 | 0.28 | 0.00 | 2.85 | 1.10 | 0.79 | 0.06 | 0.00 | 0.49 | 1.67 |
| Senior Enlisted | 12.23 | 39.15 | 19.37 | 2.78 | 18.38 | 33.92 | 145.51 | 149.93 | 26.40 | 88.94 | 53.66 | 20.85 | 127.91 | 111.96 | 27.36 | 72.02 | 2.20 | 20.32 | 8.78 | 6.64 | 9.49 | 40.75 |
| Junior Officer | 3.53 | 32.25 | 6.90 | 2.09 | 11.19 | 2.73 | 29.15 | 14.08 | 0.00 | 11.49 | 11.34 | 1.24 | 13.77 | 10.29 | 6.58 | 7.97 | 0.26 | 3.52 | 3.22 | 3.24 | 2.56 | 5.26 |
| Senior Officer | | 12.87 | 51.47 | 25.22 | 29.85 | | 0.70 | 31.89 | 24.34 | 18.98 | 24.42 | | 3.51 | 28.86 | 22.15 | 18.17 | | 0.56 | 7.58 | 6.60 | 4.91 | 11.54 |
| **Wrist Hand** | **13.46** | **13.02** | **11.87** | **3.20** | **10.18** | **11.31** | **19.54** | **18.52** | **4.55** | **13.62** | **11.90** | **10.52** | **35.23** | **31.61** | **9.86** | **22.56** | **0.70** | **3.48** | **2.26** | **1.70** | **2.12** | **12.34** |
| Junior Enlisted | 29.11 | 3.75 | 0.04 | 0.00 | 8.23 | 15.31 | 2.29 | 0.01 | 0.00 | 4.40 | 6.31 | 7.83 | 4.23 | 0.22 | 0.00 | 3.07 | 0.77 | 0.48 | 0.03 | 0.00 | 0.32 | 1.70 |
| Senior Enlisted | 9.17 | 28.17 | 15.71 | 2.08 | 13.78 | 17.37 | 64.94 | 57.86 | 10.33 | 37.63 | 25.71 | 22.88 | 123.01 | 98.77 | 19.23 | 65.98 | 1.16 | 11.52 | 4.26 | 2.81 | 4.94 | 35.46 |
| Junior Officer | 2.09 | 13.96 | 4.97 | 1.44 | 5.62 | 1.23 | 10.04 | 4.80 | 0.00 | 4.02 | 4.82 | 0.84 | 11.02 | 6.27 | 4.58 | 5.68 | 0.16 | 1.69 | 1.43 | 1.18 | 1.12 | 3.40 |
| Senior Officer | | 6.18 | 26.77 | 9.27 | 14.07 | | 0.88 | 11.41 | 7.88 | 6.72 | 10.40 | | 2.65 | 21.17 | 15.63 | 13.15 | | 0.22 | 3.31 | 2.82 | 2.11 | 7.63 |
| **Total Incidence** | **325.74** | **270.43** | **257.59** | **72.58** | **225.31** | **216.78** | **402.41** | **386.05** | **93.36** | **278.51** | **251.91** | **183.11** | **611.67** | **514.48** | **154.48** | **378.12** | **32.12** | **64.15** | **45.29** | **32.40** | **44.25** | **211.19** |

**Supplemental Table 2. Incidence of neuromusculoskeletal injury-related limited duty (per 1000 episode-years) in the US Marine Corps and US Navy from December 2016 to August 2021.**

| | US Marine Corps | | | | | | | | | | Mean Total | US Navy | | | | | | | | | | Mean Total |
|---|---|---|---|---|---|---|---|---|---|---|---|---|---|---|---|---|---|---|---|---|---|---|
| | Females | | | | Mean | Males | | | | Mean | | Females | | | | Mean | Males | | | | Mean | |
| | 18-24 | 25-34 | 35-44 | 45+ | | 18-24 | 25-34 | 35-44 | 45+ | | | 18-24 | 25-34 | 35-44 | 45+ | | 18-24 | 25-34 | 35-44 | 45+ | | |
| **Ankle Foot** | **4.91** | **4.11** | **2.62** | **0.58** | **2.93** | **3.11** | **3.05** | **1.36** | **0.43** | **1.91** | **2.42** | **1.55** | **1.46** | **1.80** | **0.43** | **1.29** | **2.57** | **1.22** | **0.76** | **0.22** | **1.10** | **1.20** |
| Junior Enlisted | 7.40 | 10.83 | 0.00 | 0.00 | 4.56 | 4.61 | 6.15 | 0.00 | 0.00 | 2.69 | 3.63 | 2.84 | 3.62 | 4.13 | 0.00 | 2.65 | 2.49 | 2.65 | 1.01 | 0.00 | 1.54 | 2.09 |
| Senior Enlisted | 5.22 | 4.27 | 2.46 | 0.00 | 2.99 | 4.72 | 4.66 | 2.68 | 1.19 | 3.31 | 3.15 | 1.83 | 2.05 | 1.65 | 1.73 | 1.82 | 1.68 | 1.74 | 1.53 | 0.61 | 1.39 | 1.60 |
| Junior Officer | 2.11 | 1.35 | 4.78 | 0.00 | 2.06 | 0.00 | 1.38 | 1.77 | 0.00 | 0.79 | 1.42 | 0.00 | 0.16 | 1.19 | 0.00 | 0.34 | 3.54 | 0.50 | 0.43 | 0.00 | 1.12 | 0.73 |
| Senior Officer | | 0.00 | 3.22 | 2.31 | 1.85 | | 0.00 | 0.99 | 0.54 | 0.51 | 1.18 | | 0.00 | 0.21 | 0.00 | 0.07 | | 0.00 | 0.06 | 0.29 | 0.12 | 0.09 |
| **Arm** | **2.49** | **0.36** | **0.00** | **0.00** | **0.59** | **4.90** | **0.80** | **0.23** | **0.72** | **1.45** | **1.02** | **0.43** | **0.86** | **0.21** | **0.00** | **0.37** | **1.55** | **0.76** | **0.18** | **52.72** | **14.62** | **7.49** |
| Junior Enlisted | 2.80 | 0.00 | 0.00 | 0.00 | 0.70 | 2.46 | 0.73 | 0.00 | 0.00 | 0.80 | 0.75 | 1.29 | 2.37 | 0.00 | 0.00 | 0.92 | 2.56 | 0.00 | 0.00 | 210.53 | 53.27 | 27.09 |
| Senior Enlisted | 4.68 | 1.43 | 0.00 | 0.00 | 1.53 | 3.07 | 2.47 | 0.93 | 1.96 | 2.11 | 1.82 | 0.00 | 1.05 | 0.83 | 0.00 | 0.47 | 2.09 | 1.43 | 0.72 | 0.35 | 1.15 | 0.81 |
| Junior Officer | 0.00 | 0.00 | 0.00 | 0.00 | 0.00 | 9.15 | 0.00 | 0.00 | 0.00 | 2.29 | 1.14 | 0.00 | 0.00 | 0.00 | 0.00 | 0.00 | 0.00 | 1.61 | 0.00 | 0.00 | 0.40 | 0.20 |
| Senior Officer | | 0.00 | 0.00 | 0.00 | 0.00 | | 0.00 | 0.00 | 0.91 | 0.30 | 0.15 | | 0.00 | 0.00 | 0.00 | 0.00 | | 0.00 | 0.00 | 0.00 | 0.00 | 0.00 |
| **Elbow** | **2.77** | **2.61** | **0.34** | **3.29** | **2.22** | **2.49** | **1.17** | **0.32** | **0.16** | **0.94** | **1.58** | **0.88** | **0.18** | **0.22** | **0.63** | **0.45** | **1.23** | **0.84** | **0.16** | **0.08** | **0.53** | **0.49** |
| Junior Enlisted | 3.63 | 9.36 | 0.00 | 0.00 | 3.25 | 2.52 | 2.66 | 0.00 | 0.00 | 1.30 | 2.27 | 0.62 | 0.00 | 0.00 | 0.00 | 0.15 | 1.65 | 1.92 | 0.00 | 0.00 | 0.89 | 0.52 |
| Senior Enlisted | 4.68 | 1.10 | 1.38 | 13.16 | 5.08 | 1.19 | 1.48 | 1.29 | 0.32 | 1.07 | 3.07 | 2.02 | 0.71 | 0.89 | 1.01 | 1.16 | 2.05 | 0.53 | 0.64 | 0.31 | 0.88 | 1.02 |
| Junior Officer | 0.00 | 0.00 | 0.00 | 0.00 | 0.00 | 3.76 | 0.53 | 0.00 | 0.00 | 1.07 | 0.54 | 0.00 | 0.00 | 0.00 | 0.00 | 0.00 | 0.00 | 0.90 | 0.00 | 0.00 | 0.23 | 0.11 |
| Senior Officer | | 0.00 | 0.00 | 0.00 | 0.00 | | 0.00 | 0.00 | 0.33 | 0.11 | 0.05 | | 0.00 | 0.00 | 1.53 | 0.51 | | 0.00 | 0.00 | 0.00 | 0.00 | 0.25 |
| **Knee** | **7.66** | **9.69** | **3.43** | **1.57** | **5.45** | **8.51** | **7.21** | **2.92** | **1.15** | **4.71** | **5.08** | **8.82** | **3.79** | **2.99** | **1.09** | **3.86** | **7.78** | **4.05** | **3.15** | **0.64** | **3.65** | **3.76** |
| Junior Enlisted | 12.53 | 18.56 | 0.00 | 0.00 | 7.77 | 11.42 | 15.47 | 0.00 | 0.00 | 6.72 | 7.25 | 7.54 | 7.22 | 6.28 | 0.00 | 5.26 | 7.97 | 8.32 | 7.99 | 0.00 | 6.07 | 5.66 |
| Senior Enlisted | 10.45 | 9.16 | 8.52 | 6.28 | 8.60 | 10.95 | 9.42 | 5.90 | 2.58 | 7.21 | 7.91 | 6.68 | 5.02 | 3.11 | 1.87 | 4.17 | 9.23 | 4.57 | 3.30 | 2.14 | 4.81 | 4.49 |
| Junior Officer | 0.00 | 11.02 | 0.00 | 0.00 | 2.76 | 3.16 | 3.95 | 3.24 | 0.00 | 2.59 | 2.67 | 12.24 | 2.92 | 1.30 | 0.00 | 4.11 | 6.16 | 1.98 | 0.92 | 0.19 | 2.31 | 3.21 |
| Senior Officer | | 0.00 | 5.22 | 0.00 | 1.74 | | 0.00 | 2.53 | 2.01 | 1.51 | 1.63 | | 0.00 | 1.29 | 2.49 | 1.26 | | 1.33 | 0.39 | 0.22 | 0.65 | 0.95 |
| **Leg** | **2.08** | **2.54** | **0.00** | **0.00** | **1.09** | **1.58** | **1.97** | **1.99** | **1.06** | **1.66** | **1.37** | **0.80** | **0.66** | **0.78** | **0.00** | **0.54** | **1.68** | **1.62** | **1.35** | **0.28** | **1.21** | **0.87** |
| Junior Enlisted | 2.98 | 4.50 | 0.00 | 0.00 | 1.87 | 2.34 | 3.73 | 0.00 | 0.00 | 1.52 | 1.69 | 0.50 | 1.48 | 2.26 | 0.00 | 1.06 | 2.00 | 2.26 | 2.43 | 0.00 | 1.67 | 1.37 |
| Senior Enlisted | 3.28 | 2.99 | 0.00 | 0.00 | 1.57 | 2.41 | 2.91 | 3.17 | 1.95 | 2.61 | 2.09 | 1.90 | 1.17 | 0.84 | 0.00 | 0.98 | 3.06 | 1.98 | 2.01 | 0.19 | 1.81 | 1.39 |
| Junior Officer | 0.00 | 2.66 | 0.00 | 0.00 | 0.67 | 0.00 | 1.24 | 1.88 | 0.00 | 0.78 | 0.72 | 0.00 | 0.00 | 0.00 | 0.00 | 0.00 | 0.00 | 0.23 | 0.97 | 0.00 | 0.30 | 0.15 |
| Senior Officer | | 0.00 | 0.00 | 0.00 | 0.00 | | 0.00 | 2.92 | 2.28 | 1.73 | 0.87 | | 0.00 | 0.00 | 0.00 | 0.00 | | 2.02 | 0.00 | 0.95 | 0.99 | 0.50 |
| **Low back** | **5.48** | **5.61** | **4.01** | **0.46** | **3.79** | **4.48** | **3.45** | **3.22** | **1.01** | **2.94** | **3.36** | **3.56** | **1.54** | **1.77** | **0.64** | **1.76** | **1.90** | **1.88** | **1.84** | **0.41** | **1.48** | **1.62** |
| Junior Enlisted | 9.89 | 10.51 | 0.00 | 0.00 | 5.10 | 6.60 | 6.91 | 6.19 | 0.00 | 4.93 | 5.01 | 2.93 | 3.46 | 2.83 | 0.00 | 2.30 | 3.18 | 4.06 | 3.72 | 0.00 | 2.74 | 2.52 |
| Senior Enlisted | 6.57 | 6.36 | 3.18 | 1.85 | 4.49 | 6.30 | 5.04 | 3.52 | 3.29 | 4.54 | 4.51 | 2.59 | 2.48 | 2.02 | 1.88 | 2.24 | 1.98 | 2.31 | 2.59 | 0.95 | 1.96 | 2.10 |
| Junior Officer | 0.00 | 2.29 | 9.23 | 0.00 | 2.88 | 0.54 | 1.36 | 1.91 | 0.00 | 0.95 | 1.92 | 5.16 | 0.23 | 1.37 | 0.39 | 1.79 | 0.54 | 0.73 | 0.55 | 0.17 | 0.50 | 1.14 |
| Senior Officer | | 3.29 | 3.64 | 0.00 | 2.31 | | 0.49 | 1.25 | 0.73 | 0.82 | 1.57 | | 0.00 | 0.85 | 0.29 | 0.38 | | 0.41 | 0.51 | 0.51 | 0.48 | 0.43 |
| **Mid back** | **2.16** | **0.63** | **0.17** | **0.00** | **0.65** | **1.19** | **0.98** | **0.27** | **0.30** | **0.65** | **0.65** | **1.35** | **0.30** | **0.03** | **0.46** | **0.48** | **0.44** | **0.21** | **0.29** | **0.12** | **0.25** | **0.37** |
| Junior Enlisted | 3.64 | 1.63 | 0.00 | 0.00 | 1.32 | 2.20 | 2.28 | 0.00 | 0.00 | 1.12 | 1.22 | 0.74 | 0.79 | 0.00 | 0.00 | 0.38 | 0.63 | 0.28 | 0.00 | 0.00 | 0.23 | 0.31 |
| Senior Enlisted | 2.85 | 0.90 | 0.67 | 0.00 | 1.10 | 1.38 | 1.46 | 0.87 | 1.21 | 1.23 | 1.17 | 0.26 | 0.42 | 0.10 | 0.78 | 0.39 | 0.69 | 0.55 | 0.93 | 0.48 | 0.66 | 0.52 |
| Junior Officer | 0.00 | 0.00 | 0.00 | 0.00 | 0.00 | 0.00 | 0.18 | 0.00 | 0.00 | 0.04 | 0.02 | 3.05 | 0.00 | 0.00 | 0.00 | 0.76 | 0.00 | 0.00 | 0.24 | 0.00 | 0.06 | 0.41 |
| Senior Officer | | 0.00 | 0.00 | 0.00 | 0.00 | | 0.00 | 0.20 | 0.00 | 0.07 | 0.03 | | 0.00 | 0.00 | 1.04 | 0.35 | | 0.00 | 0.00 | 0.00 | 0.00 | 0.17 |
| **Neck** | **1.55** | **1.47** | **0.86** | **0.94** | **1.18** | **1.35** | **1.19** | **1.40** | **1.07** | **1.25** | **1.22** | **1.30** | **1.04** | **1.98** | **0.97** | **1.33** | **0.93** | **0.72** | **1.25** | **0.54** | **0.85** | **1.09** |
| Junior Enlisted | 2.29 | 2.00 | 0.00 | 0.00 | 1.07 | 1.83 | 2.34 | 0.00 | 0.00 | 1.04 | 1.06 | 1.12 | 1.84 | 4.86 | 0.00 | 1.96 | 0.76 | 1.25 | 2.90 | 0.00 | 1.23 | 1.59 |
| Senior Enlisted | 2.37 | 3.04 | 1.25 | 0.00 | 1.66 | 2.23 | 2.09 | 2.73 | 2.41 | 2.37 | 2.01 | 2.79 | 1.90 | 0.68 | 1.99 | 1.84 | 0.46 | 1.24 | 1.47 | 1.04 | 1.05 | 1.45 |
| Junior Officer | 0.00 | 0.86 | 0.00 | 0.00 | 0.21 | 0.00 | 0.34 | 2.07 | 0.00 | 0.60 | 0.41 | 0.00 | 0.41 | 1.91 | 0.65 | 0.74 | 1.58 | 0.37 | 0.46 | 0.88 | 0.82 | 0.78 |
| Senior Officer | | 0.00 | 2.19 | 3.76 | 1.98 | | 0.00 | 0.82 | 1.87 | 0.90 | 1.44 | | 0.00 | 0.48 | 1.26 | 0.58 | | 0.00 | 0.17 | 0.23 | 0.13 | 0.36 |

| | | | | | | | | | | | | | | | | | | | | | |
|---|---|---|---|---|---|---|---|---|---|---|---|---|---|---|---|---|---|---|---|---|---|
| **Other** | **1.12** | **1.00** | **0.25** | **0.00** | **0.56** | **1.08** | **0.71** | **0.14** | **0.17** | **0.49** | **0.52** | **0.53** | **2.39** | **0.07** | **0.00** | **0.76** | **0.59** | **0.44** | **0.30** | **0.11** | **0.34** | **0.55** |
| Junior Enlisted | 0.84 | 2.77 | 0.00 | 0.00 | 0.90 | 0.83 | 0.32 | 0.00 | 0.00 | 0.29 | 0.60 | 1.04 | 1.84 | 0.00 | 0.00 | 0.72 | 1.07 | 0.97 | 0.00 | 0.00 | 0.51 | 0.61 |
| Senior Enlisted | 2.52 | 1.24 | 1.00 | 0.00 | 1.19 | 2.40 | 1.17 | 0.55 | 0.00 | 1.03 | 1.11 | 0.54 | 0.56 | 0.28 | 0.00 | 0.34 | 0.69 | 0.78 | 0.26 | 0.45 | 0.55 | 0.45 |
| Junior Officer | 0.00 | 0.00 | 0.00 | 0.00 | 0.00 | 0.00 | 1.36 | 0.00 | 0.00 | 0.34 | 0.17 | 0.00 | 1.48 | 0.00 | 0.00 | 0.37 | 0.00 | 0.00 | 0.94 | 0.00 | 0.23 | 0.30 |
| Senior Officer | | 0.00 | 0.00 | 0.00 | 0.00 | | 0.00 | 0.00 | 0.68 | 0.23 | 0.11 | | 5.69 | 0.00 | 0.00 | 1.90 | | 0.00 | 0.00 | 0.00 | 0.00 | 0.95 |
| **Pelvis Hip** | **17.66** | **12.06** | **2.79** | **0.00** | **7.49** | **6.89** | **9.26** | **4.35** | **1.36** | **5.37** | **6.43** | **7.29** | **5.41** | **3.61** | **0.77** | **4.07** | **4.89** | **5.37** | **4.73** | **0.82** | **3.89** | **3.98** |
| Junior Enlisted | 25.31 | 20.53 | 0.00 | 0.00 | 11.46 | 10.00 | 15.86 | 0.00 | 0.00 | 6.46 | 8.96 | 8.79 | 8.90 | 8.96 | 0.00 | 6.66 | 8.66 | 7.64 | 12.38 | 0.00 | 7.17 | 6.92 |
| Senior Enlisted | 16.40 | 15.77 | 6.96 | 0.00 | 9.78 | 10.67 | 9.74 | 5.49 | 3.46 | 7.34 | 8.56 | 7.23 | 5.85 | 3.51 | 1.56 | 4.54 | 6.02 | 4.53 | 3.98 | 2.30 | 4.21 | 4.37 |
| Junior Officer | 11.28 | 11.96 | 0.00 | 0.00 | 5.81 | 0.00 | 2.79 | 9.87 | 0.00 | 3.16 | 4.49 | 5.85 | 2.98 | 0.87 | 0.00 | 2.42 | 0.00 | 1.19 | 1.49 | 0.42 | 0.78 | 1.60 |
| Senior Officer | | 0.00 | 4.21 | 0.00 | 1.40 | | 8.65 | 2.05 | 1.97 | 4.23 | 2.81 | | 3.90 | 1.11 | 1.50 | 2.17 | | 8.10 | 1.07 | 0.55 | 3.24 | 2.70 |
| **Shoulder** | **6.62** | **6.25** | **3.76** | **1.95** | **4.51** | **8.18** | **10.36** | **4.07** | **2.27** | **6.09** | **5.30** | **3.66** | **2.44** | **2.96** | **1.16** | **2.48** | **6.02** | **3.04** | **3.17** | **1.05** | **3.14** | **2.81** |
| Junior Enlisted | 11.08 | 14.48 | 0.00 | 0.00 | 6.39 | 11.40 | 13.77 | 0.00 | 0.00 | 6.29 | 6.34 | 3.56 | 5.46 | 8.65 | 0.00 | 4.42 | 6.80 | 6.90 | 5.82 | 0.00 | 4.88 | 4.65 |
| Senior Enlisted | 8.77 | 9.47 | 8.06 | 3.51 | 7.45 | 10.25 | 8.54 | 7.94 | 6.52 | 8.31 | 7.88 | 2.63 | 2.57 | 2.16 | 2.01 | 2.34 | 7.96 | 3.90 | 4.13 | 2.82 | 4.70 | 3.52 |
| Junior Officer | 0.00 | 1.05 | 4.90 | 0.00 | 1.49 | 2.88 | 4.46 | 5.31 | 0.00 | 3.16 | 2.32 | 4.78 | 1.72 | 0.58 | 0.90 | 1.99 | 3.29 | 1.35 | 2.14 | 0.53 | 1.83 | 1.91 |
| Senior Officer | | 0.00 | 2.11 | 4.30 | 2.13 | | 14.69 | 3.02 | 2.54 | 6.75 | 4.44 | | 0.00 | 0.45 | 1.75 | 0.73 | | 0.00 | 0.58 | 0.85 | 0.48 | 0.61 |
| **Wrist Hand** | **2.66** | **1.64** | **0.31** | **1.17** | **1.36** | **1.11** | **1.21** | **0.47** | **0.00** | **0.67** | **1.02** | **0.73** | **1.08** | **2.44** | **0.21** | **1.14** | **2.24** | **0.85** | **1.43** | **0.07** | **1.07** | **1.11** |
| Junior Enlisted | 2.65 | 2.42 | 0.00 | 0.00 | 1.27 | 2.35 | 3.39 | 0.00 | 0.00 | 1.43 | 1.35 | 2.19 | 2.71 | 7.39 | 0.00 | 3.07 | 3.01 | 2.41 | 3.57 | 0.00 | 2.25 | 2.66 |
| Senior Enlisted | 5.32 | 4.16 | 1.24 | 4.68 | 3.85 | 0.99 | 1.06 | 1.07 | 0.00 | 0.78 | 2.31 | 0.00 | 1.07 | 0.22 | 0.00 | 0.32 | 1.00 | 0.74 | 1.28 | 0.28 | 0.83 | 0.58 |
| Junior Officer | 0.00 | 0.00 | 0.00 | 0.00 | 0.00 | 0.00 | 0.39 | 0.82 | 0.00 | 0.30 | 0.15 | 0.00 | 0.54 | 0.94 | 0.00 | 0.37 | 2.70 | 0.26 | 0.30 | 0.00 | 0.81 | 0.59 |
| Senior Officer | | 0.00 | 0.00 | 0.00 | 0.00 | | 0.00 | 0.00 | 0.00 | 0.00 | 0.00 | | 0.00 | 1.22 | 0.83 | 0.68 | | 0.00 | 0.57 | 0.00 | 0.19 | 0.44 |
| **Total Incidence** | **57.17** | **47.98** | **18.55** | **9.96** | **31.83** | **44.87** | **41.37** | **20.74** | **9.69** | **28.12** | **29.98** | **30.90** | **21.14** | **18.86** | **6.36** | **18.54** | **31.83** | **20.99** | **18.62** | **57.06** | **32.14** | **25.34** |

**Supplemental Table 3. Incidence of neuromusculoskeletal injury-related long-term disability (per 1000 episode-years) in the US Marine Corps and US Navy from December 2016 to August 2021.**

| | US Marine Corps | | | | | | | | | | Mean Total | US Navy | | | | | | | | | | Mean Total |
|---|---|---|---|---|---|---|---|---|---|---|---|---|---|---|---|---|---|---|---|---|---|---|
| | Females | | | | Mean | Males | | | | Mean | | Females | | | | Mean | Males | | | | Mean | |
| | 18-24 | 25-34 | 35-44 | 45+ | | 18-24 | 25-34 | 35-44 | 45+ | | | 18-24 | 25-34 | 35-44 | 45+ | | 18-24 | 25-34 | 35-44 | 45+ | | |
| **Ankle Foot** | **0.92** | **1.01** | **0.00** | **0.00** | **0.45** | **0.64** | **0.58** | **0.08** | **0.03** | **0.31** | **0.38** | **0.22** | **0.22** | **0.29** | **0.05** | **0.19** | **0.11** | **0.21** | **0.05** | **0.00** | **0.09** | **0.14** |
| Junior Enlisted | 1.46 | 2.89 | 0.00 | 0.00 | 1.09 | 0.93 | 1.06 | 0.00 | 0.00 | 0.50 | 0.79 | 0.22 | 0.71 | 1.03 | 0.00 | 0.49 | 0.34 | 0.42 | 0.00 | 0.00 | 0.19 | 0.34 |
| Senior Enlisted | 1.30 | 1.14 | 0.00 | 0.00 | 0.61 | 0.98 | 1.12 | 0.22 | 0.12 | 0.61 | 0.61 | 0.46 | 0.17 | 0.12 | 0.19 | 0.23 | 0.00 | 0.32 | 0.09 | 0.00 | 0.10 | 0.17 |
| Junior Officer | 0.00 | 0.00 | 0.00 | 0.00 | 0.00 | 0.00 | 0.13 | 0.00 | 0.00 | 0.03 | 0.02 | 0.00 | 0.00 | 0.00 | 0.00 | 0.00 | 0.00 | 0.10 | 0.11 | 0.00 | 0.05 | 0.03 |
| Senior Officer | | 0.00 | 0.00 | 0.00 | 0.00 | | 0.00 | 0.10 | 0.00 | 0.03 | 0.02 | | 0.00 | 0.00 | 0.00 | 0.00 | | 0.00 | 0.00 | 0.00 | 0.00 | 0.00 |
| **Arm** | **0.23** | **0.36** | **0.00** | **0.00** | **0.14** | **0.04** | **0.04** | **0.00** | **0.00** | **0.02** | **0.08** | **0.14** | **0.48** | **0.10** | **0.00** | **0.19** | **1.01** | **0.06** | **0.00** | **26.32** | **7.23** | **3.71** |
| Junior Enlisted | 0.70 | 0.00 | 0.00 | 0.00 | 0.17 | 0.11 | 0.00 | 0.00 | 0.00 | 0.03 | 0.10 | 0.43 | 1.58 | 0.00 | 0.00 | 0.50 | 0.93 | 0.00 | 0.00 | 105.26 | 26.55 | 13.53 |
| Senior Enlisted | 0.00 | 1.43 | 0.00 | 0.00 | 0.36 | 0.00 | 0.16 | 0.00 | 0.00 | 0.04 | 0.20 | 0.00 | 0.35 | 0.42 | 0.00 | 0.19 | 2.09 | 0.22 | 0.00 | 0.00 | 0.58 | 0.39 |
| Junior Officer | 0.00 | 0.00 | 0.00 | 0.00 | 0.00 | 0.00 | 0.00 | 0.00 | 0.00 | 0.00 | 0.00 | 0.00 | 0.00 | 0.00 | 0.00 | 0.00 | 0.00 | 0.00 | 0.00 | 0.00 | 0.00 | 0.00 |
| Senior Officer | | 0.00 | 0.00 | 0.00 | 0.00 | | 0.00 | 0.00 | 0.00 | 0.00 | 0.00 | | 0.00 | 0.00 | 0.00 | 0.00 | | 0.00 | 0.00 | 0.00 | 0.00 | 0.00 |
| **Elbow** | **0.30** | **1.17** | **0.00** | **0.00** | **0.37** | **0.17** | **0.03** | **0.03** | **0.00** | **0.05** | **0.21** | **0.00** | **0.00** | **0.00** | **0.38** | **0.10** | **0.53** | **0.24** | **0.00** | **0.00** | **0.17** | **0.14** |
| Junior Enlisted | 0.91 | 4.68 | 0.00 | 0.00 | 1.40 | 0.52 | 0.00 | 0.00 | 0.00 | 0.13 | 0.76 | 0.00 | 0.00 | 0.00 | 0.00 | 0.00 | 0.55 | 0.96 | 0.00 | 0.00 | 0.38 | 0.19 |
| Senior Enlisted | 0.00 | 0.00 | 0.00 | 0.00 | 0.00 | 0.00 | 0.11 | 0.14 | 0.00 | 0.06 | 0.03 | 0.00 | 0.00 | 0.00 | 0.00 | 0.00 | 1.03 | 0.00 | 0.00 | 0.00 | 0.26 | 0.13 |
| Junior Officer | 0.00 | 0.00 | 0.00 | 0.00 | 0.00 | 0.00 | 0.00 | 0.00 | 0.00 | 0.00 | 0.00 | 0.00 | 0.00 | 0.00 | 0.00 | 0.00 | 0.00 | 0.00 | 0.00 | 0.00 | 0.00 | 0.00 |
| Senior Officer | | 0.00 | 0.00 | 0.00 | 0.00 | | 0.00 | 0.00 | 0.00 | 0.00 | 0.00 | | 0.00 | 0.00 | 1.53 | 0.51 | | 0.00 | 0.00 | 0.00 | 0.00 | 0.25 |
| **Knee** | **1.94** | **1.42** | **0.00** | **0.00** | **0.77** | **1.18** | **1.10** | **0.16** | **0.00** | **0.57** | **0.67** | **1.15** | **0.47** | **0.41** | **0.08** | **0.49** | **0.76** | **0.41** | **0.23** | **0.02** | **0.33** | **0.41** |
| Junior Enlisted | 2.85 | 1.59 | 0.00 | 0.00 | 1.11 | 1.76 | 2.67 | 0.00 | 0.00 | 1.11 | 1.11 | 1.45 | 1.18 | 1.57 | 0.00 | 1.05 | 1.15 | 1.03 | 0.73 | 0.00 | 0.73 | 0.89 |
| Senior Enlisted | 2.99 | 0.78 | 0.00 | 0.00 | 0.94 | 1.78 | 1.58 | 0.52 | 0.00 | 0.97 | 0.96 | 2.01 | 0.71 | 0.07 | 0.31 | 0.77 | 1.11 | 0.59 | 0.05 | 0.07 | 0.46 | 0.62 |
| Junior Officer | 0.00 | 3.31 | 0.00 | 0.00 | 0.83 | 0.00 | 0.16 | 0.00 | 0.00 | 0.04 | 0.43 | 0.00 | 0.00 | 0.00 | 0.00 | 0.00 | 0.00 | 0.00 | 0.15 | 0.00 | 0.04 | 0.02 |
| Senior Officer | | 0.00 | 0.00 | 0.00 | 0.00 | | 0.00 | 0.13 | 0.00 | 0.04 | 0.02 | | 0.00 | 0.00 | 0.00 | 0.00 | | 0.00 | 0.00 | 0.00 | 0.00 | 0.00 |
| **Leg** | **0.54** | **0.65** | **0.00** | **0.00** | **0.28** | **0.23** | **0.29** | **0.04** | **0.00** | **0.14** | **0.21** | **0.02** | **0.16** | **0.04** | **0.00** | **0.06** | **0.31** | **0.09** | **0.37** | **0.00** | **0.18** | **0.12** |
| Junior Enlisted | 0.81 | 1.50 | 0.00 | 0.00 | 0.58 | 0.35 | 0.67 | 0.00 | 0.00 | 0.25 | 0.42 | 0.06 | 0.42 | 0.00 | 0.00 | 0.12 | 0.36 | 0.19 | 1.22 | 0.00 | 0.44 | 0.28 |
| Senior Enlisted | 0.82 | 1.12 | 0.00 | 0.00 | 0.48 | 0.34 | 0.49 | 0.17 | 0.00 | 0.25 | 0.37 | 0.00 | 0.23 | 0.17 | 0.00 | 0.10 | 0.56 | 0.19 | 0.25 | 0.00 | 0.25 | 0.17 |
| Junior Officer | 0.00 | 0.00 | 0.00 | 0.00 | 0.00 | 0.00 | 0.00 | 0.00 | 0.00 | 0.00 | 0.00 | 0.00 | 0.00 | 0.00 | 0.00 | 0.00 | 0.00 | 0.00 | 0.00 | 0.00 | 0.00 | 0.00 |
| Senior Officer | | 0.00 | 0.00 | 0.00 | 0.00 | | 0.00 | 0.00 | 0.00 | 0.00 | 0.00 | | 0.00 | 0.00 | 0.00 | 0.00 | | 0.00 | 0.00 | 0.00 | 0.00 | 0.00 |
| **Low back** | **1.15** | **0.66** | **1.11** | **0.00** | **0.70** | **0.83** | **0.83** | **0.25** | **0.00** | **0.45** | **0.58** | **0.40** | **0.27** | **0.30** | **0.05** | **0.24** | **0.40** | **0.45** | **0.22** | **0.03** | **0.26** | **0.25** |
| Junior Enlisted | 2.06 | 0.00 | 0.00 | 0.00 | 0.51 | 1.25 | 1.46 | 0.00 | 0.00 | 0.68 | 0.60 | 0.55 | 0.67 | 0.71 | 0.00 | 0.48 | 0.71 | 1.04 | 0.62 | 0.00 | 0.59 | 0.54 |
| Senior Enlisted | 1.39 | 2.25 | 0.75 | 0.00 | 1.10 | 1.25 | 1.22 | 0.49 | 0.00 | 0.74 | 0.92 | 0.65 | 0.29 | 0.32 | 0.19 | 0.36 | 0.48 | 0.47 | 0.19 | 0.07 | 0.30 | 0.33 |
| Junior Officer | 0.00 | 0.38 | 3.69 | 0.00 | 1.02 | 0.00 | 0.16 | 0.44 | 0.00 | 0.15 | 0.58 | 0.00 | 0.11 | 0.00 | 0.00 | 0.03 | 0.00 | 0.27 | 0.07 | 0.00 | 0.09 | 0.06 |
| Senior Officer | | 0.00 | 0.00 | 0.00 | 0.00 | | 0.49 | 0.05 | 0.00 | 0.18 | 0.09 | | 0.00 | 0.17 | 0.00 | 0.06 | | 0.00 | 0.00 | 0.06 | 0.02 | 0.04 |
| **Mid back** | **0.97** | **0.37** | **0.00** | **0.00** | **0.29** | **0.23** | **0.35** | **0.02** | **0.00** | **0.15** | **0.22** | **1.10** | **0.08** | **0.00** | **0.00** | **0.24** | **0.12** | **0.05** | **0.02** | **0.00** | **0.04** | **0.14** |
| Junior Enlisted | 1.21 | 0.81 | 0.00 | 0.00 | 0.51 | 0.57 | 0.86 | 0.00 | 0.00 | 0.36 | 0.43 | 0.25 | 0.34 | 0.00 | 0.00 | 0.15 | 0.20 | 0.07 | 0.00 | 0.00 | 0.07 | 0.11 |
| Senior Enlisted | 1.71 | 0.67 | 0.00 | 0.00 | 0.60 | 0.13 | 0.37 | 0.10 | 0.00 | 0.15 | 0.37 | 0.00 | 0.00 | 0.00 | 0.00 | 0.00 | 0.17 | 0.15 | 0.07 | 0.00 | 0.10 | 0.05 |
| Junior Officer | 0.00 | 0.00 | 0.00 | 0.00 | 0.00 | 0.00 | 0.18 | 0.00 | 0.00 | 0.04 | 0.02 | 3.05 | 0.00 | 0.00 | 0.00 | 0.76 | 0.00 | 0.00 | 0.00 | 0.00 | 0.00 | 0.38 |
| Senior Officer | | 0.00 | 0.00 | 0.00 | 0.00 | | 0.00 | 0.00 | 0.00 | 0.00 | 0.00 | | 0.00 | 0.00 | 0.00 | 0.00 | | 0.00 | 0.00 | 0.00 | 0.00 | 0.00 |
| **Neck** | **0.05** | **0.63** | **0.00** | **0.00** | **0.18** | **0.31** | **0.26** | **0.15** | **0.09** | **0.20** | **0.19** | **0.04** | **0.21** | **0.91** | **0.33** | **0.39** | **0.15** | **0.21** | **0.75** | **0.06** | **0.30** | **0.35** |
| Junior Enlisted | 0.16 | 1.00 | 0.00 | 0.00 | 0.29 | 0.32 | 0.64 | 0.00 | 0.00 | 0.24 | 0.26 | 0.13 | 0.66 | 3.24 | 0.00 | 1.01 | 0.23 | 0.50 | 2.90 | 0.00 | 0.91 | 0.96 |

| | | | | | | | | | | | | | | | | | | | | | |
|---|---|---|---|---|---|---|---|---|---|---|---|---|---|---|---|---|---|---|---|---|---|
| Senior Enlisted | 0.00 | 0.65 | 0.00 | 0.00 | 0.16 | 0.61 | 0.38 | 0.27 | 0.19 | 0.36 | 0.26 | 0.00 | 0.20 | 0.00 | 0.66 | 0.21 | 0.23 | 0.23 | 0.09 | 0.08 | 0.16 | 0.19 |
| Junior Officer | 0.00 | 0.86 | 0.00 | 0.00 | 0.21 | 0.00 | 0.00 | 0.35 | 0.00 | 0.09 | 0.15 | 0.00 | 0.00 | 0.38 | 0.65 | 0.26 | 0.00 | 0.12 | 0.00 | 0.18 | 0.07 | 0.17 |
| Senior Officer | | 0.00 | 0.00 | 0.00 | 0.00 | | 0.00 | 0.00 | 0.19 | 0.06 | 0.03 | | 0.00 | 0.00 | 0.00 | 0.00 | | 0.00 | 0.00 | 0.00 | 0.00 | 0.00 |
| **Other** | **0.06** | **0.45** | **0.00** | **0.00** | **0.13** | **0.39** | **0.14** | **0.00** | **0.00** | **0.11** | **0.12** | **0.03** | **0.40** | **0.00** | **0.00** | **0.11** | **0.30** | **0.19** | **0.00** | **0.00** | **0.11** | **0.11** |
| Junior Enlisted | 0.17 | 1.39 | 0.00 | 0.00 | 0.39 | 0.28 | 0.00 | 0.00 | 0.00 | 0.07 | 0.23 | 0.09 | 0.41 | 0.00 | 0.00 | 0.12 | 0.56 | 0.56 | 0.00 | 0.00 | 0.28 | 0.20 |
| Senior Enlisted | 0.00 | 0.41 | 0.00 | 0.00 | 0.10 | 0.90 | 0.55 | 0.00 | 0.00 | 0.36 | 0.23 | 0.00 | 0.19 | 0.00 | 0.00 | 0.05 | 0.35 | 0.18 | 0.00 | 0.00 | 0.13 | 0.09 |
| Junior Officer | 0.00 | 0.00 | 0.00 | 0.00 | 0.00 | 0.00 | 0.00 | 0.00 | 0.00 | 0.00 | 0.00 | 0.00 | 0.99 | 0.00 | 0.00 | 0.25 | 0.00 | 0.00 | 0.00 | 0.00 | 0.00 | 0.12 |
| Senior Officer | | 0.00 | 0.00 | 0.00 | 0.00 | | 0.00 | 0.00 | 0.00 | 0.00 | 0.00 | | 0.00 | 0.00 | 0.00 | 0.00 | | 0.00 | 0.00 | 0.00 | 0.00 | 0.00 |
| **Pelvis Hip** | **3.33** | **2.59** | **0.00** | **0.00** | **1.36** | **1.32** | **1.56** | **0.43** | **0.00** | **0.80** | **1.08** | **1.27** | **1.11** | **2.39** | **0.00** | **1.19** | **1.28** | **0.85** | **0.07** | **0.06** | **0.52** | **0.85** |
| Junior Enlisted | 5.24 | 5.25 | 0.00 | 0.00 | 2.62 | 2.18 | 4.04 | 0.00 | 0.00 | 1.55 | 2.09 | 2.36 | 2.30 | 8.96 | 0.00 | 3.40 | 2.35 | 2.15 | 0.00 | 0.00 | 1.12 | 2.26 |
| Senior Enlisted | 4.76 | 5.10 | 0.00 | 0.00 | 2.47 | 1.78 | 2.21 | 0.61 | 0.00 | 1.15 | 1.81 | 1.45 | 1.65 | 0.58 | 0.00 | 0.92 | 1.50 | 0.86 | 0.00 | 0.23 | 0.65 | 0.78 |
| Junior Officer | 0.00 | 0.00 | 0.00 | 0.00 | 0.00 | 0.00 | 0.00 | 0.82 | 0.00 | 0.21 | 0.10 | 0.00 | 0.50 | 0.00 | 0.00 | 0.12 | 0.00 | 0.40 | 0.00 | 0.00 | 0.10 | 0.11 |
| Senior Officer | | 0.00 | 0.00 | 0.00 | 0.00 | | 0.00 | 0.29 | 0.00 | 0.10 | 0.05 | | 0.00 | 0.00 | 0.00 | 0.00 | | 0.00 | 0.27 | 0.00 | 0.09 | 0.04 |
| **Shoulder** | **0.61** | **1.01** | **3.80** | **0.00** | **1.40** | **1.02** | **1.91** | **0.40** | **0.07** | **0.84** | **1.12** | **0.19** | **0.30** | **0.77** | **0.00** | **0.32** | **0.59** | **0.43** | **0.79** | **0.01** | **0.45** | **0.39** |
| Junior Enlisted | 1.82 | 2.23 | 0.00 | 0.00 | 1.01 | 1.65 | 1.44 | 0.00 | 0.00 | 0.77 | 0.89 | 0.56 | 0.87 | 2.88 | 0.00 | 1.08 | 1.06 | 0.92 | 2.91 | 0.00 | 1.22 | 1.15 |
| Senior Enlisted | 0.00 | 0.75 | 0.50 | 0.00 | 0.31 | 1.42 | 1.18 | 0.37 | 0.26 | 0.81 | 0.56 | 0.00 | 0.34 | 0.20 | 0.00 | 0.13 | 0.71 | 0.56 | 0.13 | 0.06 | 0.36 | 0.25 |
| Junior Officer | 0.00 | 1.05 | 14.69 | 0.00 | 3.93 | 0.00 | 0.14 | 1.12 | 0.00 | 0.31 | 2.12 | 0.00 | 0.00 | 0.00 | 0.00 | 0.00 | 0.00 | 0.24 | 0.13 | 0.00 | 0.09 | 0.05 |
| Senior Officer | | 0.00 | 0.00 | 0.00 | 0.00 | | 4.90 | 0.11 | 0.00 | 1.67 | 0.83 | | 0.00 | 0.00 | 0.00 | 0.00 | | 0.00 | 0.00 | 0.00 | 0.00 | 0.00 |
| **Wrist Hand** | **0.92** | **0.65** | **0.00** | **0.00** | **0.36** | **0.05** | **0.21** | **0.00** | **0.00** | **0.07** | **0.21** | **0.24** | **0.22** | **0.92** | **0.00** | **0.35** | **0.41** | **0.21** | **0.54** | **0.00** | **0.29** | **0.32** |
| Junior Enlisted | 0.62 | 1.21 | 0.00 | 0.00 | 0.46 | 0.15 | 0.80 | 0.00 | 0.00 | 0.24 | 0.35 | 0.73 | 0.77 | 3.69 | 0.00 | 1.30 | 0.57 | 0.69 | 1.78 | 0.00 | 0.76 | 1.03 |
| Senior Enlisted | 2.13 | 1.39 | 0.00 | 0.00 | 0.88 | 0.00 | 0.05 | 0.00 | 0.00 | 0.01 | 0.45 | 0.00 | 0.09 | 0.00 | 0.00 | 0.02 | 0.67 | 0.17 | 0.09 | 0.00 | 0.23 | 0.13 |
| Junior Officer | 0.00 | 0.00 | 0.00 | 0.00 | 0.00 | 0.00 | 0.00 | 0.00 | 0.00 | 0.00 | 0.00 | 0.00 | 0.00 | 0.00 | 0.00 | 0.00 | 0.00 | 0.00 | 0.30 | 0.00 | 0.08 | 0.04 |
| Senior Officer | | 0.00 | 0.00 | 0.00 | 0.00 | | 0.00 | 0.00 | 0.00 | 0.00 | 0.00 | | 0.00 | 0.00 | 0.00 | 0.00 | | 0.00 | 0.00 | 0.00 | 0.00 | 0.00 |
| **Total Incidence** | **11.04** | **10.96** | **4.91** | **0.00** | **6.44** | **6.42** | **7.29** | **1.57** | **0.19** | **3.70** | **5.07** | **4.81** | **3.93** | **6.13** | **0.88** | **3.88** | **5.97** | **3.40** | **3.04** | **26.50** | **9.98** | **6.93** |

Supplemental Table 4. Results of the hurdle negative binomial regression assessing body region, sex, age, rank, and service branch on risk of neuromusculoskeletal injury episodes (adjusted for zero deflation) in US Navy (USN) and US Marine Corps personnel.

| | **US Navy** | | | | | | **US Marine Corps** | | | | | | **Total** | | | | | |
|---|---|---|---|---|---|---|---|---|---|---|---|---|---|---|---|---|---|---|
| | Zero Model | | | Count model | | | Zero Model | | | Count model | | | Zero Model | | | Count model | | |
| | OR | 95% CI | | RR | 95% CI | | OR | 95% CI | | RR | 95% CI | | OR | 95% CI | | RR | 95% CI | |
| | | LL | UL | | LL | UL | | LL | UL | | LL | UL | | LL | UL | | LL | UL |
| **Region**: Arm | **0.12** | **0.02** | **0.62** | 1.10 | 0.52 | 2.32 | 0.12 | 0.00 | 7.96 | **0.09** | **0.05** | **0.17** | **0.22** | **0.07** | **0.77** | **0.20** | **0.12** | **0.33** |
| Elbow | **0.16** | **0.03** | **0.80** | **0.15** | **0.08** | **0.28** | 1.00 | 0.01 | 95.26 | **0.16** | **0.09** | **0.29** | 0.31 | 0.09 | 1.08 | **0.16** | **0.10** | **0.26** |
| Knee | 0.68 | 0.12 | 3.80 | 0.60 | 0.34 | 1.06 | 1.00 | 0.01 | 95.26 | 0.71 | 0.41 | 1.23 | 0.80 | 0.22 | 2.95 | 0.66 | 0.42 | 1.04 |
| Leg | 0.49 | 0.09 | 2.61 | **0.27** | **0.15** | **0.49** | 0.12 | 0.00 | 7.96 | **0.46** | **0.26** | **0.80** | 0.54 | 0.15 | 1.91 | **0.37** | **0.23** | **0.59** |
| Low back | 1.57 | 0.24 | 10.13 | 1.52 | 0.88 | 2.62 | 1.00 | 0.01 | 95.26 | 1.63 | 0.95 | 2.80 | 1.27 | 0.33 | 4.87 | **1.57** | **1.00** | **2.45** |
| Mid back | 0.68 | 0.12 | 3.80 | **0.46** | **0.26** | **0.81** | 0.12 | 0.00 | 7.96 | **0.49** | **0.28** | **0.84** | 0.65 | 0.18 | 2.36 | **0.47** | **0.30** | **0.74** |
| Neck | 1.00 | 0.17 | 5.90 | 0.64 | 0.36 | 1.11 | 1.00 | 0.01 | 95.26 | **0.58** | **0.34** | **1.00** | 1.00 | 0.27 | 3.75 | **0.61** | **0.39** | **0.96** |
| Pelvis-hip | 0.27 | 0.05 | 1.38 | **0.27** | **0.15** | **0.49** | 1.00 | 0.01 | 95.26 | **0.49** | **0.29** | **0.85** | 0.44 | 0.13 | 1.57 | **0.37** | **0.23** | **0.58** |
| Shoulder | 0.68 | 0.12 | 3.80 | **0.54** | **0.30** | **0.96** | 1.00 | 0.01 | 95.26 | 0.64 | 0.37 | 1.11 | 0.80 | 0.22 | 2.95 | **0.59** | **0.38** | **0.94** |
| Wrist & Hand | 0.36 | 0.07 | 1.87 | **0.33** | **0.18** | **0.60** | 1.00 | 0.01 | 95.26 | **0.29** | **0.17** | **0.50** | 0.54 | 0.15 | 1.91 | **0.31** | **0.20** | **0.50** |
| Other | 0.36 | 0.07 | 1.87 | **0.28** | **0.15** | **0.50** | 1.00 | 0.01 | 95.26 | **0.22** | **0.13** | **0.39** | 0.54 | 0.15 | 1.91 | **0.25** | **0.16** | **0.40** |
| **Sex:** Males | **0.21** | **0.10** | **0.43** | **0.13** | **0.10** | **0.18** | **0.02** | **0.00** | **0.23** | 1.05 | 0.81 | 1.35 | **0.27** | **0.16** | **0.46** | **0.40** | **0.32** | **0.50** |
| **Age Range**: | | | | | | | | | | | | | | | | | | |
| 25-34 | 2.12 | 0.77 | 5.83 | **2.01** | **1.25** | **3.24** | $34.61*10^{6}$ | 0.00 | ∞ | 1.38 | 0.92 | 2.05 | 1.68 | 0.71 | 4.00 | **1.57** | **1.08** | **2.28** |
| 35-44 | 0.97 | 0.37 | 2.50 | **1.97** | **1.18** | **3.30** | **0.00** | **0.00** | **0.00** | 1.48 | 0.94 | 2.32 | **0.19** | **0.09** | **0.41** | **1.66** | **1.11** | **2.50** |
| 45+ | 0.61 | 0.24 | 1.55 | 1.13 | 0.65 | 1.94 | **0.00** | **0.00** | **0.00** | **0.45** | **0.27** | **0.75** | **0.07** | **0.03** | **0.16** | 0.68 | 0.44 | 1.06 |
| **Rank:** Senior Enlisted | **102.13** | **21.69** | **480.86** | **4.66** | **2.75** | **7.91** | $0.13*10^{15}$ | 0.00 | ∞ | 1.53 | 0.96 | 2.43 | **$0.29*10^{3}$** | **64.36** | **$1.33*10^{3}$** | **2.64** | **1.74** | **4.01** |
| Junior Officers | **10.56** | **4.69** | **23.78** | 0.57 | 0.32 | 1.01 | **$14.08*10^{3}$** | **$0.21*10^{3}$** | **$0.95*10^{6}$** | **0.37** | **0.23** | **0.59** | **12.47** | **6.64** | **23.43** | **0.38** | **0.25** | **0.58** |
| Senior Officers | **15.18** | **5.68** | **40.62** | 1.54 | 0.83 | 2.87 | $0.20*10^{15}$ | 0.00 | ∞ | 1.08 | 0.63 | 1.88 | **61.27** | **24.39** | **153.95** | 0.97 | 0.60 | 1.58 |
| **Branch:** USN | **-** | **-** | **-** | **-** | **-** | **-** | **-** | **-** | **-** | **-** | **-** | **-** | 0.67 | 0.41 | 1.12 | **0.48** | **0.39** | **0.60** |
| Negative Log-Likelihood: | 1011 on 39 *Df* | | | | | | 1141 on 39 *Df* | | | | | | 2284 on 41 *Df* | | | | | |

Bolded figures depict statistical significance ($p$<0.05); OR, odds ratio; RR, rate ratio; CI, confidence interval.
Contrasts: Body Region [ankle-foot]; Sex [Female]; Age [18-24]; Rank [Junior Enlisted]; Branch [US Marine Corps]

Supplemental Table 5. Results of the hurdle negative binomial regression assessing body region, sex, age, rank, and service branch on risk of neuromusculoskeletal injury-related episodes of limited duty (adjusted for zero deflation) in US Navy (USN) and US Marine Corps personnel.

| | **US Navy** | | | | | | **US Marine Corps** | | | | | | **Total** | | | | | |
|---|---|---|---|---|---|---|---|---|---|---|---|---|---|---|---|---|---|---|
| | Zero Model | | | Count model | | | Zero Model | | | Count model | | | Zero Model | | | Count model | | |
| | | 95% CI | | | 95% CI | | | 95% CI | | | 95% CI | | | 95% CI | | | 95% CI | |
| | OR | LL | UL | RR | LL | UL | OR | LL | UL | RR | LL | UL | OR | LL | UL | RR | LL | UL |
| **Body Region**: | | | | | | | | | | | | | | | | | | |
| Arm | **0.10** | **0.02** | **0.38** | **9.61** | **4.99** | **18.49** | **0.13** | **0.03** | **0.51** | 0.84 | 0.55 | 1.29 | **2.65** | **1.10** | **6.36** | **4.48** | **2.89** | **6.94** |
| Elbow | **0.15** | **0.04** | **0.57** | 0.77 | 0.45 | 1.32 | 0.32 | 0.08 | 1.22 | 0.77 | 0.53 | 1.12 | **0.11** | **0.04** | **0.30** | 0.77 | 0.54 | 1.10 |
| Knee | 3.16 | 0.68 | 14.58 | **2.71** | **1.74** | **4.22** | 0.79 | 0.20 | 3.04 | **2.20** | **1.56** | **3.11** | **0.22** | **0.09** | **0.57** | **2.46** | **1.80** | **3.35** |
| Leg | 0.34 | 0.09 | 1.26 | 1.05 | 0.63 | 1.72 | 0.32 | 0.08 | 1.22 | 0.88 | 0.61 | 1.28 | 1.44 | 0.55 | 3.79 | 0.96 | 0.68 | 1.35 |
| Low back | 4.72 | 0.92 | 24.27 | 1.17 | 0.76 | 1.82 | **2.17** | **0.52** | **8.98** | 1.24 | 0.89 | 1.72 | 0.34 | 0.13 | 0.86 | 1.19 | 0.88 | 1.61 |
| Mid back | **0.23** | **0.06** | **0.85** | **0.52** | **0.31** | **0.87** | **0.20** | **0.05** | **0.79** | **0.36** | **0.24** | **0.53** | **2.92** | **1.04** | **8.24** | **0.42** | **0.29** | **0.60** |
| Neck | 2.25 | 0.52 | 9.71 | 0.93 | 0.59 | 1.45 | 0.63 | 0.16 | 2.40 | **0.67** | **0.47** | **0.96** | **0.22** | **0.09** | **0.57** | 0.76 | 0.56 | 1.05 |
| Pelvis-hip | 3.16 | 0.68 | 14.58 | **2.96** | **1.89** | **4.62** | 0.79 | 0.20 | 3.04 | **2.82** | **1.99** | **3.98** | 1.13 | 0.43 | 2.91 | **2.87** | **2.10** | **3.92** |
| Shoulder | 3.16 | 0.68 | 14.58 | **2.09** | **1.35** | **3.26** | 1.65 | 0.41 | 6.66 | **2.29** | **1.63** | **3.20** | 1.44 | 0.55 | 3.79 | **2.17** | **1.60** | **2.95** |
| Wrist & Hand | 0.64 | 0.17 | 2.39 | 0.96 | 0.60 | 1.55 | **0.20** | **0.05** | **0.79** | **0.56** | **0.38** | **0.82** | 2.16 | 0.79 | 5.87 | 0.77 | 0.55 | 1.08 |
| Other | 0.19 | 0.05 | 0.70 | 0.91 | 0.52 | 1.57 | **0.16** | **0.04** | **0.63** | **0.35** | **0.23** | **0.52** | **0.38** | **0.15** | **0.95** | **0.60** | **0.42** | **0.87** |
| **Sex:** Males | **1.99** | **1.11** | **3.58** | 0.83 | 0.67 | 1.02 | **3.49** | **1.96** | **6.20** | **0.67** | **0.57** | **0.78** | **0.18** | **0.07** | **0.46** | **0.75** | **0.65** | **0.87** |
| **Age Range**: 25-34 | 0.87 | 0.35 | 2.20 | **0.68** | **0.51** | **0.91** | 0.76 | 0.32 | 1.81 | 0.86 | 0.71 | 1.05 | **2.55** | **1.71** | **3.80** | **0.82** | **0.68** | **1.00** |
| 35-44 | 0.95 | 0.37 | 2.39 | **0.59** | **0.45** | **0.78** | **0.19** | **0.08** | **0.45** | **0.71** | **0.56** | **0.90** | 0.82 | 0.44 | 1.53 | **0.72** | **0.60** | **0.88** |
| 45+ | **0.13** | **0.05** | **0.33** | **0.60** | **0.42** | **0.86** | **0.04** | **0.02** | **0.12** | **0.74** | **0.56** | **0.98** | **0.41** | **0.22** | **0.75** | 0.83 | 0.65 | 1.07 |
| **Rank:** | | | | | | | | | | | | | | | | | | |
| Senior Enlisted | **16.41** | **5.50** | **48.93** | **0.46** | **0.35** | **0.60** | **19.59** | **7.58** | **50.66** | **0.75** | **0.61** | **0.93** | **0.08** | **0.04** | **0.16** | **0.54** | **0.47** | **0.63** |
| Junior Officers | 0.55 | 0.27 | 1.13 | **0.34** | **0.25** | **0.46** | **0.46** | **0.23** | **0.94** | **0.54** | **0.42** | **0.70** | **2.27** | **1.53** | **3.38** | **0.51** | **0.42** | **0.61** |
| Senior Officers | **0.42** | **0.19** | **0.93** | **0.33** | **0.23** | **0.48** | 1.04 | 0.48 | 2.26 | **0.46** | **0.34** | **0.62** | **16.12** | **8.05** | **32.30** | **0.38** | **0.31** | **0.47** |
| **Branch:** USN | - | - | - | - | - | - | - | - | - | - | - | - | **0.52** | **0.32** | **0.86** | **0.34** | **0.27** | **0.44** |
| Negative Log-Likelihood: | 1649 on 39 *Df* | | | | | | 1466 on 39 *Df* | | | | | | 3179 on 41 *Df* | | | | | |

Bolded figures depict statistical significance ($p<0.05$); OR, odds ratio; RR, rate ratio; CI, confidence interval.
Contrasts: Body Region [ankle-foot]; Sex [Female]; Age [18-24]; Rank [Junior Enlisted]; Branch [US Marine Corps]

Supplemental Table 6. Results of the hurdle negative binomial regression assessing body region, sex, age, rank, and service branch on risk of neuromusculoskeletal injury-related episodes of long-term disability (adjusted for zero deflation) in US Navy (USN) and US Marine Corps personnel.

| | **US Navy** | | | | | | **US Marine Corps** | | | | | | **Total** | | | | | |
|---|---|---|---|---|---|---|---|---|---|---|---|---|---|---|---|---|---|---|
| | Zero Model | | | Count model | | | Zero Model | | | Count model | | | Zero Model | | | Count model | | |
| | | 95% CI | | | 95% CI | | | 95% CI | | | 95% CI | | | 95% CI | | | 95% CI | |
| | OR | LL | UL | RR | LL | UL | OR | LL | UL | RR | LL | UL | OR | LL | UL | RR | LL | UL |
| **Body Region**: | | | | | | | | | | | | | | | | | | |
| Arm | 0.26 | 0.06 | 1.12 | **20.04** | **7.16** | **56.09** | **0.07** | **0.01** | **0.40** | **0.43** | **0.19** | **0.96** | **0.16** | **0.06** | **0.47** | **14.27** | **5.88** | **34.61** |
| Elbow | **0.07** | **0.01** | **0.37** | **4.56** | **1.44** | **14.43** | **0.11** | **0.02** | **0.57** | 0.86 | 0.40 | 1.83 | **0.10** | **0.03** | **0.30** | 1.83 | 0.88 | 3.80 |
| Knee | 1.69 | 0.41 | 7.03 | **2.57** | **1.31** | **5.04** | 1.00 | 0.23 | 4.35 | 1.72 | 0.97 | 3.03 | 1.29 | 0.48 | 3.49 | **2.10** | **1.26** | **3.50** |
| Leg | 0.45 | 0.11 | 1.90 | 1.26 | 0.59 | 2.69 | 0.42 | 0.09 | 1.88 | 0.64 | 0.35 | 1.18 | 0.45 | 0.16 | 1.25 | 0.97 | 0.56 | 1.71 |
| Low back | **4.91** | **1.14** | **21.23** | 1.42 | 0.74 | 2.70 | 2.30 | 0.53 | 10.05 | 1.36 | 0.78 | 2.37 | **3.15** | **1.15** | **8.57** | 1.35 | 0.83 | 2.20 |
| Mid back | 0.26 | 0.06 | 1.12 | 1.68 | 0.69 | 4.10 | 0.56 | 0.13 | 2.50 | 0.60 | 0.33 | 1.09 | 0.39 | 0.14 | 1.09 | 0.96 | 0.53 | 1.73 |
| Neck | 2.20 | 0.53 | 9.18 | 1.55 | 0.79 | 3.06 | 1.00 | 0.23 | 4.35 | 0.68 | 0.37 | 1.23 | 1.47 | 0.54 | 3.97 | 1.04 | 0.61 | 1.74 |
| Pelvis-hip | 1.30 | 0.31 | 5.40 | **5.26** | **2.67** | **10.38** | 0.75 | 0.17 | 3.30 | **2.95** | **1.65** | **5.26** | 1.00 | 0.37 | 2.71 | **3.89** | **2.32** | **6.53** |
| Shoulder | 1.30 | 0.31 | 5.40 | **2.24** | **1.12** | **4.44** | 3.05 | 0.69 | 13.39 | **2.75** | **1.54** | **4.93** | 1.89 | 0.70 | 5.11 | **2.67** | **1.58** | **4.51** |
| Wrist & Hand | 0.59 | 0.14 | 2.46 | 1.97 | 0.94 | 4.10 | 0.22 | 0.05 | 1.05 | 0.66 | 0.34 | 1.29 | 0.39 | 0.14 | 1.09 | 1.47 | 0.83 | 2.60 |
| Other | 0.26 | 0.06 | 1.12 | 1.57 | 0.69 | 3.54 | **0.16** | **0.03** | **0.78** | 0.64 | 0.32 | 1.30 | **0.22** | **0.08** | **0.63** | 1.00 | 0.54 | 1.86 |
| **Sex:** Males | **2.08** | **1.14** | **3.81** | **0.73** | **0.53** | **1.00** | **3.23** | **1.67** | **6.22** | **0.39** | **0.29** | **0.52** | **2.44** | **1.59** | **3.76** | **0.57** | **0.45** | **0.73** |
| **Age Range**: | | | | | | | | | | | | | | | | | | |
| 25-34 | **2.41** | **1.02** | **5.68** | **0.65** | **0.44** | **0.97** | 1.81 | 0.78 | 4.22 | 1.08 | 0.80 | 1.46 | **2.06** | **1.14** | **3.73** | 0.82 | 0.62 | 1.10 |
| 35-44 | 0.70 | 0.31 | 1.59 | 0.89 | 0.56 | 1.43 | **0.17** | **0.07** | **0.41** | **0.53** | **0.33** | **0.87** | **0.36** | **0.20** | **0.64** | 0.89 | 0.62 | 1.27 |
| 45+ | **0.09** | **0.04** | **0.23** | 1.34 | 0.62 | 2.91 | **0.01** | **0.00** | **0.05** | **0.37** | **0.15** | **0.87** | **0.05** | **0.02** | **0.10** | 1.36 | 0.67 | 2.76 |
| **Rank:** | | | | | | | | | | | | | | | | | | |
| Senior Enlisted | 1.07 | 0.52 | 2.20 | **0.32** | **0.22** | **0.45** | **2.21** | **1.00** | **4.91** | 0.82 | 0.60 | 1.11 | **1.62** | **1.06** | **2.48** | **0.52** | **0.41** | **0.67** |
| Junior Officers | **0.05** | **0.02** | **0.13** | **0.41** | **0.24** | **0.72** | **0.09** | **0.04** | **0.22** | 1.02 | 0.56 | 1.84 | 1.46 | 0.87 | 2.45 | **0.43** | **0.33** | **0.57** |
| Senior Officers | **0.01** | **0.00** | **0.04** | **0.16** | **0.05** | **0.49** | **0.07** | **0.02** | **0.21** | 0.60 | 0.32 | 1.12 | **0.08** | **0.04** | **0.14** | **0.58** | **0.39** | **0.87** |
| **Branch:** USN | - | - | - | - | - | - | - | - | - | - | - | - | **0.03** | **0.01** | **0.07** | **0.35** | **0.19** | **0.66** |
| Negative Log-Likelihood: | 857 on 39 *Df* | | | | | | 767 on 39 *Df* | | | | | | 1679 on 41 *Df* | | | | | |

Bolded figures depict statistical significance ($p<0.05$); OR, odds ratio; RR, rate ratio; CI, confidence interval.
Contrasts: Body Region [ankle-foot]; Sex [Female]; Age [18-24]; Rank [Junior Enlisted]; Branch [US Marine Corps]

Supplemental Table 7. Comparison of the results of the negative binomial regression models (with and without adjustment for zero deflation) assessing body region, sex, age, rank, and service branch on risk on episodes of neuromusculoskeletal injury in US Navy (USN) and US Marine Corps personnel.

| | **US Navy** | | | | | | **US Marine Corps** | | | | | | **Total** | | | | | |
|---|---|---|---|---|---|---|---|---|---|---|---|---|---|---|---|---|---|---|
| | Hurdle Model | | | Standard Model | | | Hurdle Model | | | Standard Model | | | Hurdle Model | | | Standard Model | | |
| | RR | 95% CI | | RR | 95% CI | | RR | 95% CI | | RR | 95% CI | | RR | 95% CI | | RR | 95% CI | |
| | | LL | UL | | LL | UL | | LL | UL | | LL | UL | | LL | UL | | LL | UL |
| **Region**: Arm | 1.10 | 0.52 | 2.32 | 0.77 | 0.42 | 1.44 | **0.09** | **0.05** | **0.17** | **0.12** | **0.07** | **0.22** | **0.20** | **0.12** | **0.33** | **0.23** | **0.14** | **0.36** |
| Elbow | **0.15** | **0.08** | **0.28** | **0.17** | **0.09** | **0.30** | **0.16** | **0.09** | **0.29** | **0.20** | **0.11** | **0.37** | **0.16** | **0.10** | **0.26** | **0.19** | **0.12** | **0.29** |
| Knee | 0.60 | 0.34 | 1.06 | 0.62 | 0.37 | 1.06 | 0.71 | 0.41 | 1.23 | 0.74 | 0.42 | 1.29 | 0.66 | 0.42 | 1.04 | **0.69** | **0.45** | **1.05** |
| Leg | **0.27** | **0.15** | **0.49** | **0.32** | **0.18** | **0.55** | **0.46** | **0.26** | **0.80** | **0.46** | **0.26** | **0.80** | **0.37** | **0.23** | **0.59** | **0.39** | **0.26** | **0.60** |
| Low back | 1.52 | 0.88 | 2.62 | 1.47 | 0.88 | 2.47 | 1.63 | 0.95 | 2.80 | 1.56 | 0.90 | 2.72 | **1.57** | **1.00** | **2.45** | **1.52** | **1.00** | **2.31** |
| Mid back | **0.46** | **0.26** | **0.81** | **0.49** | **0.29** | **0.84** | **0.49** | **0.28** | **0.84** | **0.47** | **0.27** | **0.82** | **0.47** | **0.30** | **0.74** | **0.47** | **0.31** | **0.73** |
| Neck | 0.64 | 0.36 | 1.11 | 0.67 | 0.40 | 1.14 | **0.58** | **0.34** | **1.00** | 0.59 | 0.34 | 1.04 | **0.61** | **0.39** | **0.96** | **0.63** | **0.41** | **0.96** |
| Pelvis-hip | **0.27** | **0.15** | **0.49** | **0.29** | **0.17** | **0.50** | **0.49** | **0.29** | **0.85** | **0.53** | **0.30** | **0.94** | **0.37** | **0.23** | **0.58** | **0.39** | **0.26** | **0.61** |
| Shoulder | **0.54** | **0.30** | **0.96** | **0.55** | **0.32** | **0.95** | 0.64 | 0.37 | 1.11 | 0.66 | 0.38 | 1.17 | **0.59** | **0.38** | **0.94** | **0.61** | **0.40** | **0.93** |
| Wrist & Hand | **0.33** | **0.18** | **0.60** | **0.35** | **0.20** | **0.61** | **0.29** | **0.17** | **0.50** | **0.32** | **0.18** | **0.57** | **0.31** | **0.20** | **0.50** | **0.34** | **0.22** | **0.52** |
| Other | **0.28** | **0.15** | **0.50** | **0.31** | **0.18** | **0.53** | **0.22** | **0.13** | **0.39** | **0.25** | **0.14** | **0.44** | **0.25** | **0.16** | **0.40** | **0.28** | **0.18** | **0.43** |
| **Sex:** Males | **0.13** | **0.10** | **0.18** | **0.17** | **0.13** | **0.21** | 1.05 | 0.81 | 1.35 | 0.96 | 0.74 | 1.24 | **0.40** | **0.32** | **0.50** | **0.41** | **0.34** | **0.50** |
| **Age Range**: | | | | | | | | | | | | | | | | | | |
| 25-34 | **2.01** | **1.25** | **3.24** | 1.09 | 0.73 | 1.63 | 1.38 | 0.92 | 2.05 | **0.50** | **0.32** | **0.77** | **1.57** | **1.08** | **2.28** | **0.61** | **0.44** | **0.85** |
| 35-44 | **1.97** | **1.18** | **3.30** | 0.90 | 0.59 | 1.38 | 1.48 | 0.94 | 2.32 | **0.29** | **0.18** | **0.47** | **1.66** | **1.11** | **2.50** | **0.44** | **0.31** | **0.63** |
| 45+ | 1.13 | 0.65 | 1.94 | **0.52** | **0.33** | **0.80** | **0.45** | **0.27** | **0.75** | **0.08** | **0.05** | **0.12** | 0.68 | 0.44 | 1.06 | **0.17** | **0.12** | **0.25** |
| **Rank:** Senior Enlisted | **4.66** | **2.75** | **7.91** | **11.22** | **7.66** | **16.43** | 1.53 | 0.96 | 2.43 | **8.26** | **5.34** | **12.79** | **2.64** | **1.74** | **4.01** | **11.24** | **8.16** | **15.48** |
| Junior Officers | 0.57 | 0.32 | 1.01 | **1.70** | **1.14** | **2.53** | **0.37** | **0.23** | **0.59** | **1.57** | **1.03** | **2.40** | **0.38** | **0.25** | **0.58** | **1.71** | **1.24** | **2.36** |
| Senior Officers | 1.54 | 0.83 | 2.87 | **4.23** | **2.72** | **6.60** | 1.08 | 0.63 | 1.88 | **8.51** | **5.24** | **13.87** | 0.97 | 0.60 | 1.58 | **5.61** | **3.90** | **8.09** |
| **Branch:** USN | - | - | - | - | - | - | - | - | - | - | - | - | **0.48** | **0.39** | **0.60** | **0.53** | **0.44** | **0.65** |
| Negative Log-Likelihood: | 1011 on 39 *Df* | | | 2090 on 39 *Df* | | | 1141 on 39 *Df* | | | 2508 on 39 *Df* | | | 2284 on 41 *Df* | | | 4739 on 41 *Df* | | |

Bolded figures depict statistical significance ($p$<0.05); OR, odds ratio; RR, rate ratio; CI, confidence interval.
Contrasts: Body Region [ankle-foot]; Sex [Female]; Age [18-24]; Rank [Junior Enlisted]; Branch [US Marine Corps]

Supplemental Table 8. Comparison of the results of the negative binomial regression models (with and without adjustment for zero deflation) assessing body region, sex, age, rank, and service branch on risk of episodes of neuromusculoskeletal injury-related limited duty in US Navy (USN) and US Marine Corps personnel.

| | **US Navy** | | | | | | **US Marine Corps** | | | | | | **Total** | | | | | |
|---|---|---|---|---|---|---|---|---|---|---|---|---|---|---|---|---|---|---|
| | Hurdle Model | | | Standard Model | | | Hurdle Model | | | Standard Model | | | Hurdle Model | | | Standard Model | | |
| | RR | 95% CI LL | 95% CI UL | RR | 95% CI LL | 95% CI UL | RR | 95% CI LL | 95% CI UL | RR | 95% CI LL | 95% CI UL | RR | 95% CI LL | 95% CI UL | RR | 95% CI LL | 95% CI UL |
| **Region**: Arm | **9.61** | **4.99** | **18.49** | * | * | * | 0.84 | 0.55 | 1.29 | * | * | * | **4.48** | **2.89** | **6.94** | **2.99** | **1.10** | **8.16** |
| Elbow | 0.77 | 0.45 | 1.32 | * | * | * | 0.77 | 0.53 | 1.12 | * | * | * | 0.77 | 0.54 | 1.10 | 0.51 | 0.22 | 1.18 |
| Knee | **2.71** | **1.74** | **4.22** | * | * | * | **2.20** | **1.56** | **3.11** | * | * | * | **2.46** | **1.80** | **3.35** | **2.38** | **1.04** | **5.42** |
| Leg | 1.05 | 0.63 | 1.72 | * | * | * | 0.88 | 0.61 | 1.28 | * | * | * | 0.96 | 0.68 | 1.35 | 0.63 | 0.28 | 1.46 |
| Low back | 1.17 | 0.76 | 1.82 | * | * | * | 1.24 | 0.89 | 1.72 | * | * | * | 1.19 | 0.88 | 1.61 | 1.43 | 0.63 | 3.24 |
| Mid back | **0.52** | **0.31** | **0.87** | * | * | * | **0.36** | **0.24** | **0.53** | * | * | * | **0.42** | **0.29** | **0.60** | **0.25** | **0.11** | **0.57** |
| Neck | 0.93 | 0.59 | 1.45 | * | * | * | **0.67** | **0.47** | **0.96** | * | * | * | 0.76 | 0.56 | 1.05 | 0.87 | 0.38 | 1.99 |
| Pelvis-hip | **2.96** | **1.89** | **4.62** | * | * | * | **2.82** | **1.99** | **3.98** | * | * | * | **2.87** | **2.10** | **3.92** | **2.79** | **1.22** | **6.35** |
| Shoulder | **2.09** | **1.35** | **3.26** | * | * | * | **2.29** | **1.63** | **3.20** | * | * | * | **2.17** | **1.60** | **2.95** | **2.40** | **1.05** | **5.47** |
| Wrist & Hand | 0.96 | 0.60 | 1.55 | * | * | * | **0.56** | **0.38** | **0.82** | * | * | * | 0.77 | 0.55 | 1.08 | 0.54 | 0.24 | 1.24 |
| Other | 0.91 | 0.52 | 1.57 | * | * | * | **0.35** | **0.23** | **0.52** | * | * | * | **0.60** | **0.42** | **0.87** | **0.31** | **0.13** | **0.72** |
| **Sex:** Males | 0.83 | 0.67 | 1.02 | * | * | * | **0.67** | **0.57** | **0.78** | * | * | * | **0.75** | **0.65** | **0.87** | 1.00 | 0.70 | 1.42 |
| **Age Range**: | | | | | | | | | | | | | | | | | | |
| 25-34 | **0.68** | **0.51** | **0.91** | * | * | * | 0.86 | 0.71 | 1.05 | * | * | * | **0.82** | **0.68** | **1.00** | 0.82 | 0.49 | 1.35 |
| 35-44 | **0.59** | **0.45** | **0.78** | * | * | * | **0.71** | **0.56** | **0.90** | * | * | * | **0.72** | **0.60** | **0.88** | **0.55** | **0.33** | **0.91** |
| 45+ | **0.60** | **0.42** | **0.86** | * | * | * | **0.74** | **0.56** | **0.98** | * | * | * | 0.83 | 0.65 | 1.07 | **0.36** | **0.20** | **0.65** |
| **Rank:** Senior Enlisted | **0.46** | **0.35** | **0.60** | * | * | * | **0.75** | **0.61** | **0.93** | * | * | * | **0.54** | **0.47** | **0.63** | 0.90 | 0.54 | 1.50 |
| Junior Officers | **0.34** | **0.25** | **0.46** | * | * | * | **0.54** | **0.42** | **0.70** | * | * | * | **0.51** | **0.42** | **0.61** | **0.27** | **0.17** | **0.44** |
| Senior Officers | **0.33** | **0.23** | **0.48** | * | * | * | **0.46** | **0.34** | **0.62** | * | * | * | **0.38** | **0.31** | **0.47** | **0.35** | **0.19** | **0.63** |
| **Branch:** USN | - | - | - | - | - | - | - | - | - | - | - | - | **0.34** | **0.27** | **0.44** | 0.81 | 0.57 | 1.16 |
| Negative Log-Likelihood: | 1649 on 39 *Df* | | | | | | 1466 on 39 *Df* | | | | | | 3179 on 41 *Df* | | | 7255 on 41 *Df* | | |

Bolded figures depict statistical significance ($p<0.05$); OR, odds ratio; RR, rate ratio; CI, confidence interval.
Contrasts: Body Region [ankle-foot]; Sex [Female]; Age [18-24]; Rank [Junior Enlisted]; Branch [US Marine Corps]
* Standard binomial regression model would not converge without adjustment.

Supplemental Table 9. Comparison of the results of the negative binomial regression models (with and without adjustment for zero deflation) assessing body region, sex, age, rank, and service branch on risk of neuromusculoskeletal injury-related episodes of long-term disability in US Navy (USN) and US Marine Corps personnel.

| | **US Navy** | | | | | | **US Marine Corps** | | | | | | **Total** | | | | | |
|---|---|---|---|---|---|---|---|---|---|---|---|---|---|---|---|---|---|---|
| | Hurdle Model | | | Standard Model | | | Hurdle Model | | | Standard Model | | | Hurdle Model | | | Standard Model | | |
| | RR | 95% CI | | RR | 95% CI | | RR | 95% CI | | RR | 95% CI | | RR | 95% CI | | RR | 95% CI | |
| | | LL | UL | | LL | UL | | LL | UL | | LL | UL | | LL | UL | | LL | UL |
| **Region**: Arm | **20.04** | **7.16** | **56.09** | * | * | * | **0.43** | **0.19** | **0.96** | * | * | * | **14.27** | **5.88** | **34.61** | * | * | * |
| Elbow | **4.56** | **1.44** | **14.43** | * | * | * | 0.86 | 0.40 | 1.83 | * | * | * | 1.83 | 0.88 | 3.80 | * | * | * |
| Knee | **2.57** | **1.31** | **5.04** | * | * | * | 1.72 | 0.97 | 3.03 | * | * | * | **2.10** | **1.26** | **3.50** | * | * | * |
| Leg | 1.26 | 0.59 | 2.69 | * | * | * | 0.64 | 0.35 | 1.18 | * | * | * | 0.97 | 0.56 | 1.71 | * | * | * |
| Low back | 1.42 | 0.74 | 2.70 | * | * | * | 1.36 | 0.78 | 2.37 | * | * | * | 1.35 | 0.83 | 2.20 | * | * | * |
| Mid back | 1.68 | 0.69 | 4.10 | * | * | * | 0.60 | 0.33 | 1.09 | * | * | * | 0.96 | 0.53 | 1.73 | * | * | * |
| Neck | 1.55 | 0.79 | 3.06 | * | * | * | 0.68 | 0.37 | 1.23 | * | * | * | 1.04 | 0.61 | 1.74 | * | * | * |
| Pelvis-hip | **5.26** | **2.67** | **10.38** | * | * | * | **2.95** | **1.65** | **5.26** | * | * | * | **3.89** | **2.32** | **6.53** | * | * | * |
| Shoulder | **2.24** | **1.12** | **4.44** | * | * | * | **2.75** | **1.54** | **4.93** | * | * | * | **2.67** | **1.58** | **4.51** | * | * | * |
| Wrist & Hand | 1.97 | 0.94 | 4.10 | * | * | * | 0.66 | 0.34 | 1.29 | * | * | * | 1.47 | 0.83 | 2.60 | * | * | * |
| Other | 1.57 | 0.69 | 3.54 | * | * | * | 0.64 | 0.32 | 1.30 | * | * | * | 1.00 | 0.54 | 1.86 | * | * | * |
| **Sex:** Males | **0.73** | **0.53** | **1.00** | * | * | * | **0.39** | **0.29** | **0.52** | * | * | * | **0.57** | **0.45** | **0.73** | * | * | * |
| **Age Range**: | | | | | | | | | | | | | | | | | | |
| 25-34 | **0.65** | **0.44** | **0.97** | * | * | * | 1.08 | 0.80 | 1.46 | * | * | * | 0.82 | 0.62 | 1.10 | * | * | * |
| 35-44 | 0.89 | 0.56 | 1.43 | * | * | * | **0.53** | **0.33** | **0.87** | * | * | * | 0.89 | 0.62 | 1.27 | * | * | * |
| 45+ | 1.34 | 0.62 | 2.91 | * | * | * | **0.37** | **0.15** | **0.87** | * | * | * | 1.36 | 0.67 | 2.76 | * | * | * |
| **Rank:** Senior Enlisted | **0.32** | **0.22** | **0.45** | * | * | * | 0.82 | 0.60 | 1.11 | * | * | * | **0.52** | **0.41** | **0.67** | * | * | * |
| Junior Officers | **0.41** | **0.24** | **0.72** | * | * | * | 1.02 | 0.56 | 1.84 | * | * | * | **0.43** | **0.33** | **0.57** | * | * | * |
| Senior Officers | **0.16** | **0.05** | **0.49** | * | * | * | 0.60 | 0.32 | 1.12 | * | * | * | **0.58** | **0.39** | **0.87** | * | * | * |
| **Branch:** USN | - | - | - | - | - | - | - | - | - | - | - | - | **0.35** | **0.19** | **0.66** | - | - | - |
| Negative Log-Likelihood: | 857 on 39 *Df* | | | | | | 767 on 39 *Df* | | | | | | 1679 on 41 *Df* | | | | | |

Bolded figures depict statistical significance ($p$<0.05); OR, odds ratio; RR, rate ratio; CI, confidence interval.
Contrasts: Body Region [ankle-foot]; Sex [Female]; Age [18-24]; Rank [Junior Enlisted]; Branch [US Marine Corps]
* Standard binomial regression model would not converge without adjustment.
